# Risk factors associated with SARS-CoV-2 infection and outbreaks in Long Term Care Facilities in England: a national survey

**DOI:** 10.1101/2020.10.02.20205591

**Authors:** Laura Shallcross, Danielle Burke, Owen Abbott C Stat, Alasdair Donaldson, Gemma Hallatt, Andrew Hayward, Susan Hopkins, Maria Krutikov, Katie Sharp, Leone Wardman, Sapphira Thorne

## Abstract

**Background:** Outbreaks of SARS-CoV-2 have occurred worldwide in Long Term Care Facilities (LTCFs), but the reasons why some facilities are particularly vulnerable to infection are poorly understood. We aimed to identify risk factors for SARS-CoV-2 infection and outbreaks in LTCFs.

**Methods:** Cross-sectional survey of all LTCFs providing dementia care or care to adults >65 years in England with linkage to SARS-CoV-2 test results. Exposures included: LTCF characteristics, staffing factors, and use of disease control measures. Main outcomes included risk factors for infection and outbreaks, estimated using multivariable logistic regression, and survey and test-based weighted estimates of SARS-CoV-2 prevalence.

**Findings:** 5126/9081 (56%) LTCFs participated in the survey, with 160,033 residents and 248,594 staff. The weighted period prevalence of infection in residents and staff respectively was 10.5% (95% CI: 9.9-11.1%) and 3.8% (95%: 3.4-4.2%) and 2724 LTCFs (53.1%) had ≥1 infection. Odds of infection and/or outbreaks were reduced in LTCFs that paid sickness pay, cohorted staff, did not employ agency staff and had higher staff to resident ratios. Higher odds of infection and outbreaks were identified in facilities with more admissions, lower cleaning frequency, poor compliance with isolation and “for profit” status.

**Interpretation:** Half of LTCFs had no cases suggesting they remain vulnerable to outbreaks. Reducing transmission from staff requires adequate sick pay, minimal use of temporary staff, improved staffing ratios and staff cohorting. Transmission from residents is associated with the number of admissions to the facility and poor compliance with isolation.

**Funding:** UK Government Department of Health & Social Care

**Research in context:** *Evidence before this study:* COVID-19 outbreaks have occurred worldwide in long-term care facilities (LTCFs), which provide care to elderly and vulnerable residents, and are associated with high mortality. The reasons why LTCFs are particularly vulnerable to COVID-19 are poorly understood. Most studies of risk factors for COVID-19 to date have been limited by scale, and poor quality administrative, demographic and infection control data. We conducted a systematic search on 27 July 2020 in MEDLINE Ovid, WHO COVID-19 database and in MedRxiv to identify studies reporting risk factors for COVID-19 infection or outbreaks in LTCFs, with no date or language restrictions. We used the search terms “COVID-19”, “SARS-CoV-2”, “coronavirus” and “care home”, “nursing home”, “long term care facilit” and excluded studies that did not investigate LTCF-level risk factors. 14 studies met our inclusion criteria comprising 11 cross-sectional studies and 3 surveys. The largest cross-sectional study was conducted in 9395 specialised nursing facilities across 30 states in USA; the largest survey was conducted in 124 LTCFs in Haute-Garrone region of France. Risk of bias was high across all studies, and results could not be pooled due to heterogeneity between studies. Main risk factors for infection and/or outbreaks related to the size of the facility, lower ratios of staff to residents, urban location, higher occupancy, and the community prevalence of infection. Only one study collected data on the use of disease control measures during the pandemic, and no studies provided data on risk factors such as the use of temporary staff, or the impact of staff working across multiple locations.

*Added value of this study:* We conducted a national telephone survey with managers of all LTCFs in England which provided dementia care or care to residents aged > 65 years to collect data on the number of staff and residents in each facility, confirmed SARS-CoV-2 infections, characteristics of the facility e.g.size, staffing (use of temporary staff, staffing ratios, sickness pay) and disease control measures such as cohorting and isolation. We identified risk factors for infection in residents and staff, outbreaks (defined as ≥1 case per LTCF) and large outbreaks using logistic regression. We also estimated the proportion of staff and residents who had been infected with SARS-CoV-2. Responses were obtained from 5126 of out 9081 (56%) of eligible LTCFs. To our knowledge, this is the largest and most detailed survey of risk factors for SARS-CoV-2 infection and outbreaks that has been conducted in LTCFs.

*Implications of all the available evidence:* Almost half of LTCFs surveyed in this study did not report any cases of infection, and remain vulnerable to infection and outbreaks, highlighting the need for effective control measures. Reducing transmission from staff requires adequate sick pay, minimal use of temporary staff, improved staffing ratios and staff cohorting. Transmission from residents is associated with the number of admissions to the facility and poor compliance with control measures such as isolation.

## Introduction

The global burden of SARS-CoV-2 (COVID-19) is increasing with many countries that successfully curtailed the first wave of the pandemic reporting new infections following the relaxation of lockdown measures.^1^

Long-term care facilities (LTCFs), which provide care to the elderly and those with disabilities have experienced among the highest rates of SARS-CoV-2 infection, and account for 30-50% of all COVID-19 related deaths in countries including the USA,^2^ England,^3^ Scotland,^4^ France, Spain, and Sweden.^5^ Residents of LTCFs who become infected with SARS-CoV-2 are at increased risk of severe outcomes due to their age and high prevalence of comorbidity,^6^ but they are also highly exposed to infection through frequent close contact with other residents and staff in the care setting.^7^ Undetected infection is likely to have played a key role in rapid transmission of SARS-CoV-2 in LTCFs, due to presymptomatic and asymptomatic infections and limited testing for infection at the start of the pandemic.^8,9^

In the UK, there are an estimated 400,000 residents living in approximately 11,000 LTCFs for the elderly,^10^ but there is poor quality demographic, infection prevention or administrative data available for residents and staff in these facilities. Studies based on administrative data from Canada, the USA, and Europe have identified risk factors for SARS-CoV-2 infection and outbreaks in LTCFs including: lower staff to resident ratios,^11^ increased facility size,^12^ lower care quality ratings,^11^ minority ethnic groups,^13^ and LTCFs that are “for profit” rather than state funded.^14^ However, as many studies are small and have relied on administrative data rather than surveys, they have been unable to capture the wide range of potential influences on infection.

To inform the SARS-CoV-2 pandemic response, we invited all managers of LTCFs in England to participate in a survey to collect information on the number of confirmed infections in staff and residents, staffing practices (such as employment of temporary staff and sick pay policies), use of disease control measures (such as cleaning, cohorting and isolation), and number of staff and residents. Responses were linked to results from the national testing programme. Our objectives were to identify risk factors for SARS-CoV-2 and outbreaks in residents and staff. We also estimated the prevalence of laboratory-confirmed infection in residents and staff, and the proportion of LTCFs with outbreaks.

## Methods

### Study design, setting and participants

Cross-sectional survey of managers of LTCFs in England between 26 May and 19 June 2020. Survey responses were linked to individual-level SARS-CoV-2 reverse transcription polymerase chain reaction (RT-PCR) test results obtained between 30 April and 13 June 2020 through the national testing programme which aimed to test all residents and staff.

LTCFs were eligible for the study if they mainly provided care for residents with dementia, or to adults aged > 65 years and were based in England (*N* = 9081). Eligible LTCFs were identified from a directory maintained by LangBuisson.^10^

### Survey design and telephone interviews

This survey collected data on LTCF characteristics, use of disease control measures, and the number of confirmed cases of infection in staff and residents per LTCF.

Candidate risk factors for infection were identified from the published literature and from prior knowledge of disease control measures.^15^ Experts from the Office for National Statistics designed and piloted the questionnaire using cognitive-interview methods to test comprehension, accuracy, and question acceptability.^16^ Researchers from Ipsos MORI attempted to contact 8634 (95%) of eligible LTCFs at least once and 68% of LTCFs were telephoned three or more times. Incentives to participate were not provided. Subject to informed consent, the finalised 30 minute questionnaire was delivered by telephone. Survey responses were recorded electronically and transferred securely to the NHS Covid-19 data store which is powered by Palantir Foundry.^17^

### Data Linkage

A LTCF identifier was used to attribute individual results to specific LTCFs and to link test results to survey responses, Figure 1 and supplementary material (eMethods).

**Figure 1:**
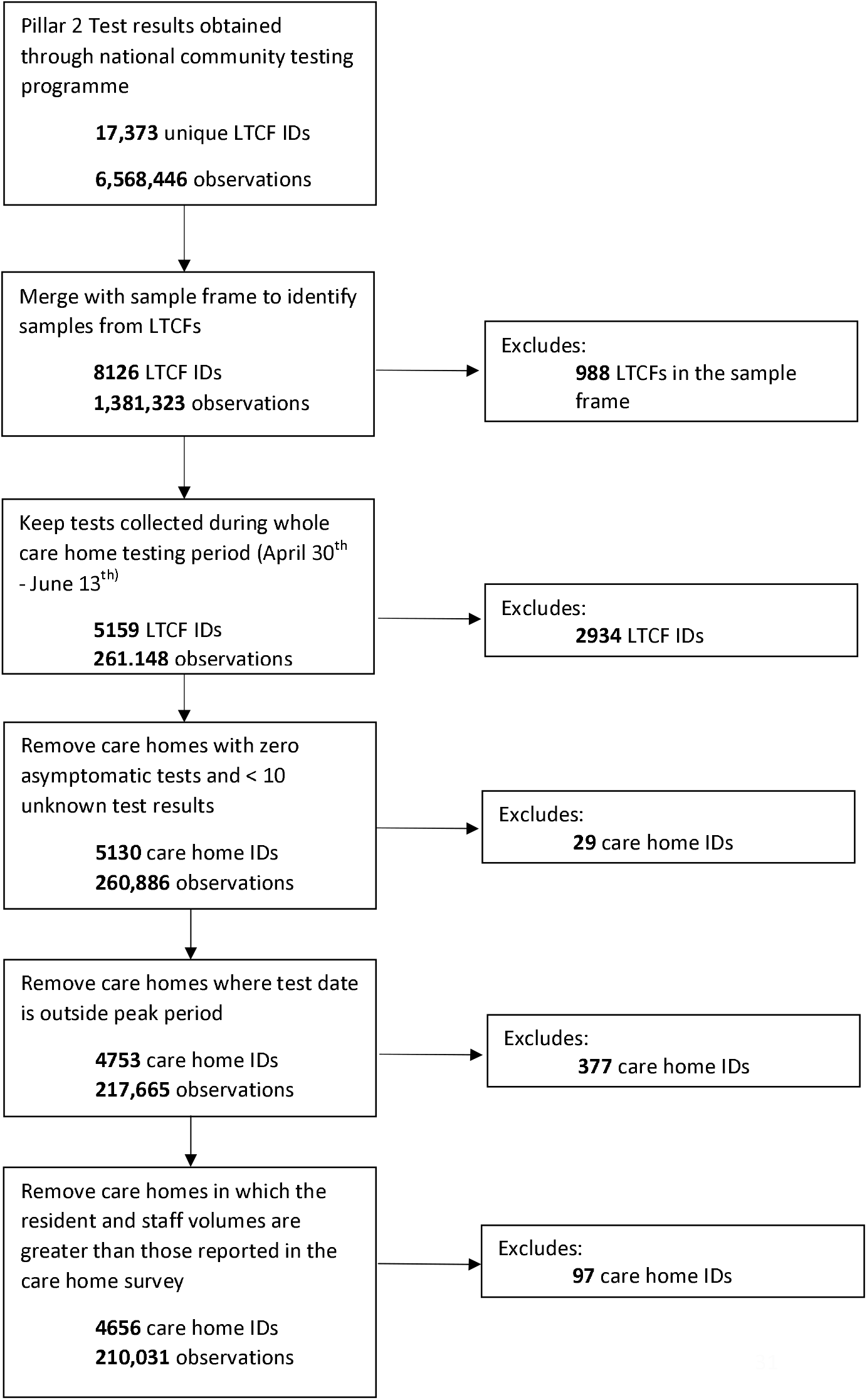
Study flow chart. The study flow chart illustrates linkage of test results from residents and staff in LTCFs to the national survey

### Exposures, outcomes, and covariates

The primary outcome was the weighted period prevalence of confirmed SARS-CoV-2 in residents and staff reported via the survey. A survey-based outcome was preferred as the primary outcome because it was not possible to reliably attribute test results to LTCFs using routine data before May 2020. Also, the testing programme measured infection at a point in time rather than over the first wave of the pandemic. There were three secondary outcomes. First, the weighted point prevalence of SARS-CoV-2 in residents and staff measured by the national testing programme (May-June 2020). Second, the proportion of care homes with an outbreak (≥1 case). Third, the proportion of LTCFs that had a large outbreak, defined as a LTCF in which at least one-third of residents and staff combined became infected with SARS-CoV-2, or the total cases in residents and staff combined was more than 20.

Care home characteristics included: Postal-code based index of multiple deprivation (IMD) and region, size, membership of a chain, funding (profit versus not for profit), and staff to bed ratio (total number of staff divided by the number of beds). In England, the quality of a LTCF is assessed across five domains by the Care Quality Commission (CQC). We included the domain “well led” in view of its relevance to the management of the pandemic. Age and sex were available for individuals tested in the national programme.

Disease control measures included: use of barrier nursing; difficulties isolating residents due to non-compliance e.g dementia; cohorting of staff with either infected or uninfected residents; cleaning frequency; payment of sickness pay to staff; use of agency (temporary staff); use of PPE; and how often staff worked at different locations. Factors were selected based on theoretical assumptions about risk factors for transmission of SARS-CoV-2 and national guidance.^18^ Week of closure to visitors was included as a continuous variable and calculated as the number of weeks since March 1^st^ 2020.

### PCR testing

PCR testing was undertaken on clinical isolates from nasopharyngeal swabs by the National Bioresource Centre using an Applied Biosytems 7500 Fast real time PCR system and the Applied Biosystems TaqPath™ 1-Step Multiplex Master Mix (No ROX) (Cat. A28523) and TaqPath COVID-19-ASY-KIT 1000 (Cat. A47817). Primer sequence details are not currently published.

### Statistical analysis

Estimates of period prevalence derived from the survey and point prevalence derived from the test data were weighted to provide nationally relevant estimates taking account of non-response by post-stratification, considering the LTCF size, IMD, and the number of LTCF run by the provider^19^.

We investigated potential risk factors for ingress of infection, and spread of infection. Multivariable logistic regression models were fitted to identify risk factors for infection in staff and residents using data from the survey and from the linked survey-test dataset. Here the outcomes of interest were binomial counts of infected residents and staff by LTCF (survey data) and presence or absence of infection (survey-test dataset). A multi-level model was fitted in the survey-test dataset to estimate individual-level and LTCF-level risk factors for infection, using random effects to account for clustering by LTCF. Multivariable logistic regression models were also fitted to identify risk factors for outbreaks (≥1 case per LTCF) and for large versus small outbreaks using survey data.

All pre-defined risk factors and potential confounding factors were included in the models unless there was strong evidence of collinearity assessed by variance inflation factors (VIF), or variables were uninformative (e.g. staff training – as almost all participants reported staff being trained). Risk factors with a VIF of >10 were excluded from the analysis. Weighting factors from the estimation of period prevalence were included in all models as adjustment factors to adjust for survey non-response.

Results were presented as proportions and/or adjusted odds ratios (aOR) with 95% confidence intervals. All reported model p-values were considered statistically significant when <0.008 (α=0.05/6) using the conservative Bonferroni adjustment for multiple hypothesis testing conducted for the six dependent variables. A heatmap was used to visualise the strength of evidence for each risk factor across all outcomes.

### Multiple imputation

Sensitivity analyses were conducted to compare complete case data (primary analysis) with models fitted to imputed data.

All analyses were undertaken using SparkR in the secure NHS Covid-19 data store.

### Ethical approval

Ethical approval was obtained from Public Health England’s Research Ethics and Governance Group reference: NR0210.

## Results

Of the 9081 eligible LTCFs in England, 5125 (56·4%) participated in the survey, providing data on 160,033 residents and 248,594 staff. Survey respondents were representative of LTCFs in England in terms of size, funding, geography, deprivation, and levels of infection, with survey weights varying from 1·5 to 2·2. A confirmed infection was reported in the survey for 19,571 residents and 10,630 staff, equivalent to a weighted period prevalence of SARS-CoV-2 of 10·5% (95% CI: 9·9-11·1%) in residents and 3·8% (95%CI: 3·4-4·2%) in staff. Of 5126 LTCFs, 2724 (53·1%) reported at least one case of SARS-CoV-2 during this period, and 469 reported a large outbreak. The weighted prevalence of infection was 2·8% (95% CI: 2·4-3·1%) in residents and 0·6% (95% CI: 0·5-0·8%) in staff, based on tests from 108,289 residents and 91,424 staff in 4656 LTCFs between April 30 – June 7. 312/323 (96·6%) of staff and 1257/1628 (77·2%) of residents with a positive test had no symptoms at the time of testing, (eTable1).

### Characteristics of LTCFs

The mean number of residents and staff per LTCF was 32·2 (SD = 17·3) and 48·5 (SD = 31·2) respectively (Table 1) and most LTCFs (4289/5126) were “For profit”. Half of LTCFs (2573; 50·2%) were single institutions and 1102 (21·5%) primarily provided dementia care. Uptake of disease control measures by care home have been previously reported.^19^

**Table 1.** Baseline characteristics of Long Term Care Facilities (LTCFs)

| **Characteristics and risk factors for infection** | **All LTCFs** |
| --- | --- |
| **Social deprivation** |  |
| Q1 (most deprived quintile) | 871 (17.0%) |
| All others | 4254 (83.0%) |
| **Provider type** |  |
| Profit | 4289 (83.7%) |
| Not for profit | 837 (16.3%) |
| **Provider size** |  |
| Single LTCF | 2573 (50.2%) |
| 2-9 LTCFs | 1471 (28.7%) |
| >10 LTCFs | 1081 (21.1%) |
| **Number of Beds** |  |
| < 25 | 1096 (21.4%) |
| 25-50 | 2497 (48.7%) |
| >50 | 1533 (29.9%) |
| **Mean number of residents (SD)** | 32.2 (17.3) |
| **Mean number of all staff (SD)** | 48.5 (31.2) |
| Mean number of nurses or carers (SD) | 33.7 (23.4) |
| Mean number of Cleaning/maintenance/catering staff (SD) | 10.4 (7.1) |
| Mean number of office or other staff (SD) | 4.5 (5.0) |
| **Region** |  |
| East Midlands | 488 (9.5%) |
| East of England | 556 (10.9%) |
| London | 282 (5.5%) |
| North East | 273 (5.3%) |
| North West | 715 (14.0%) |
| South East | 1006 (19.6%) |
| South West | 719 (14.0%) |
| West Midlands | 562 (11.0%) |
| Yorkshire and the Humber | 523 (10.2%) |
| **CQC Rating – Well led** |  |
| Outstanding/Good | 3724 (72.6%) |
| Requires improvement/Inadequate | 1323 (25.8%) |
| No rating/Not inspected | 79 (1.5%) |
| **Type of care** |  |
| Dementia | 1102 (21.5%) |
| > 65 years | 4024 (78.5%) |

Note – Denominator for LTCFs with zero cases and LTCFs with one or more cases does not include LTCFs with missing data for total number of positive cases.

### Structural factors associated with infection and outbreaks

For profit status was associated with increased odds of infection in residents (aOR 1.19, 95% CI: 1·12-1.26, p<0.001) and staff (aOR: 1·19, 95% CI: 1·10-1·28, p<0.001), Table 2, with moderate evidence of an association with large outbreaks (aOR 1·65, 95% CI: 1·07-2·54, p=0·02), Table 3. Larger care homes (>50 beds) were more likely to have ≥1 case compared with care homes that had <25 beds (aOR 2·76, 95% CI: 1·97-3·88, p<0.001), but were not more likely to have large outbreaks, and had lower odds of infection in residents (aOR 0·87, 95% CI: 0·79-0·96, p<0.001) and staff (0·79, 95% CI: 0·70-0·90, p<0.001). There was substantial regional variation in odds of infection and outbreaks in residents and staff. Residents in LTCFs with the highest levels of social deprivation had increased odds of infection (most deprived quintile versus all other quintiles: aOR: 1·08, 95% CI: 1·03-1·14, p=0·001), but outbreaks were not associated with increased deprivation.

**Table 2.** Risk Factors for infection in residents and staff of Long Term Care Facilities (LTCFs) - survey data as the outcome

|  | Residents |  |  | Staff |  |  |
| --- | --- | --- | --- | --- | --- | --- |
| Risk Factor | Proportion with infection (%) | Adjusted OR (95% CI) | p-value | Proportion with infection (%) | Adjusted OR (95% CI) | p-value |
| **Social deprivation** |  |  |  |  |  |  |
| All others | 9941 / 91048 (10.9%) | 1 | NA | 5582 / 132334 (4.2%) | 1 | NA |
| Most socially deprived quintile | 3054 / 22112 (13.8%) | 1.084 [1.032, 1.139] | 0.001 | 1469 / 31497 (4.7%) | 0.853 [0.798, 0.911] | <0.001 |
| **Care sector** |  |  |  |  |  |  |
| Not for profit | 2043 / 18860 (10.8%) | 1 | NA | 1164 / 31544 (3.7%) | 1 | NA |
| For profit | 10952 / 94300 (11.6%) | 1.187 [1.115, 1.263] | <0.001 | 5887 / 132287 (4.5%) | 1.19 [1.101, 1.288] | <0.001 |
| **Number of LTCFs in group** |  |  |  |  |  |  |
| Single | 4939 / 47282 (10.4%) | 1 | NA | 2660 / 67667 (3.9%) | 1 | NA |
| 2 - 9 | 3881 / 32992 (11.8%) | 0.983 [0.937, 1.031] | 0.482 | 2216 / 49096 (4.5%) | 0.985 [0.927, 1.047] | 0.624 |
| 10 or more | 4175 / 32886 (12.7%) | 1.039 [0.988, 1.093] | 0.140 | 2175 / 47068 (4.6%) | 0.986 [0.923, 1.052] | 0.666 |
| **Staff to bed ratio** |  |  |  |  |  |  |
| Baseline | NA | 1 | NA | NA | 1 | NA |
| One unit increased in staff to bed ratio | 12995 / 113160 (11.5%) | 0.823 [0.784, 0.865] | <0.001 | 7051 / 163831 (4.3%) | 0.632 [0.592, 0.675] | <0.001 |
| **Region** |  |  |  |  |  |  |
| London | 783 / 6368 (12.3%) | 1 | NA | 307 / 10013 (3.1%) | 1 | NA |
| East midlands | 1027 / 10255 (10.0%) | 1.021 [0.922, 1.132] | 0.685 | 620 / 14270 (4.3%) | 1.684 [1.46, 1.942] | <0.001 |
| East of England | 1320 / 13524 (9.8%) | 1.051 [0.953, 1.16] | 0.318 | 675 / 18019 (3.7%) | 1.469 [1.276, 1.69] | <0.001 |
| North East | 1259 / 7851 (16.0%) | 1.419 [1.28, 1.572] | <0.001 | 867 / 10556 (8.2%) | 3.041 [2.643, 3.499] | <0.001 |
| North West | 1918 / 15432 (12.4%) | 1.068 [0.972, 1.173] | 0.174 | 1006 / 21740 (4.6%) | 1.599 [1.397, 1.829] | <0.001 |
| South East | 1923 / 19023 (10.1%) | 0.982 [0.895, 1.076] | 0.691 | 1181 / 28408 (4.2%) | 1.5 [1.316, 1.709] | <0.001 |
| South West | 1110 / 14424 (7.7%) | 0.964 [0.871, 1.066] | 0.472 | 571 / 21573 (2.6%) | 1.243 [1.077, 1.436] | 0.003 |
| West midlands | 2205 / 14283 (15.4%) | 1.39 [1.268, 1.523] | <0.001 | 823 / 22046 (3.7%) | 1.396 [1.217, 1.601] | <0.001 |
| Yorkshire & Humber | 1450 / 12000 (12.1%) | 1.096 [0.994, 1.209] | 0.066 | 1001 / 17206 (5.8%) | 2.168 [1.893, 2.482] | <0.001 |
| **Number of care home beds** |  |  |  |  |  |  |
| Less than 25 | 580 / 8895 (6.5%) | 1 | NA | 295 / 11727 (2.5%) | 1 | NA |
| 25 to 50 | 5385 / 51225 (10.5%) | 0.948 [0.862, 1.043] | 0.271 | 2975 / 71708 (4.1%) | 0.921 [0.812, 1.044] | 0.199 |
| More than 50 | 7030 / 53040 (13.3%) | 0.873 [0.792, 0.961] | 0.006 | 3781 / 80396 (4.7%) | 0.792 [0.698, 0.9] | <0.001 |
| **Quality rating on leadership** |  |  |  |  |  |  |
| Outstanding or good | 9267 / 83987 (11.0%) | 1 | NA | 5256 / 120856 (4.3%) | 1 | NA |
| Risk Factor | Proportion with infection (%) | Adjusted OR (95% CI) | p-value | Proportion with infection (%) | Adjusted OR (95% CI) | p-value |
| Inadequate or requires improvement | 3587 / 27880 (12.9%) | 1.003 [0.96, 1.049] | 0.890 | 1689 / 40383 (4.2%) | 0.824 [0.777, 0.874] | <0.001 |
| No rating | 141 / 1293 (10.9%) | 1.03 [0.857, 1.239] | 0.750 | 106 / 2592 (4.1%) | 1.072 [0.875, 1.312] | 0.502 |
| **Main type of care provided** |  |  |  |  |  |  |
| Residents aged > 65 years | 9384 / 83998 (11.2%) | 1 | NA | 4962 / 121140 (4.1%) | 1 | NA |
| Residents with dementia | 3611 / 29162 (12.4%) | 1 | 0.113 | 2089 / 42691 (4.9%) | 1 | 0.352 |
| **Sickness payments** |  |  |  |  |  |  |
| None | 1073 / 8647 (12.4%) | 1 | NA | 681 / 13007 (5.2%) | 1 | NA |
| Statutory | 9786 / 86123 (11.4%) | 0.804 [0.749, 0.864] | <0.001 | 5112 / 120169 (4.3%) | 0.704 [0.646, 0.766] | <0.001 |
| Full or more than statutory | 2136 / 18390 (11.6%) | 0.779 [0.715, 0.849] | <0.001 | 1258 / 30655 (4.1%) | 0.679 [0.612, 0.754] | <0.001 |
| **Employment of agency nurses or carers** |  |  |  |  |  |  |
| Not employed | 3333 / 46217 (7.2%) | 1 | NA | 1657 / 62133 (2.7%) | 1 | NA |
| A few times a month | 2947 / 23299 (12.6%) | 1.566 [1.475, 1.661] | <0.001 | 1357 / 27569 (4.9%) | 1.505 [1.392, 1.627] | <0.001 |
| A few times a week | 2509 / 18082 (13.9%) | 1.338 [1.265, 1.415] | <0.001 | 1575 / 35103 (4.5%) | 1.284 [1.192, 1.383] | <0.001 |
| Most days or everyday | 4206 / 25562 (16.5%) | 1.649 [1.562, 1.741] | <0.001 | 2462 / 39026 (6.3%) | 1.849 [1.724, 1.984] | <0.001 |
| **Employment of “other” agency staff** |  |  |  |  |  |  |
| Not employed | 9129 / 87490 (10.4%) | 1 | NA | 4798 / 124394 (3.9%) | 1 | NA |
| Few times a month | 1468 / 8892 (16.5%) | 1.279 [1.199, 1.365] | <0.001 | 802 / 13553 (5.9%) | 1.306 [1.203, 1.418] | <0.001 |
| Few times a week | 1198 / 9093 (13.2%) | 1.084 [1.012, 1.162] | 0.022 | 788 / 13865 (5.7%) | 1.337 [1.231, 1.452] | <0.001 |
| Most days or everyday | 1200 / 7685 (15.6%) | 1.076 [1.002, 1.155] | 0.044 | 663 / 12019 (5.5%) | 1.037 [0.948, 1.134] | 0.424 |
| **How often staff work at other locations** |  |  |  |  |  |  |
| Not at all | 11325 / 100485 (11.3%) | 1 | NA | 5983 / 143533 (4.2%) | 1 | NA |
| Few times a month | 1086 / 7647 (14.2%) | 1.116 [1.04, 1.198] | 0.002 | 591 / 12292 (4.8%) | 1.029 [0.941, 1.126] | 0.530 |
| Few times a week | 537 / 4656 (11.5%) | 0.97 [0.882, 1.068] | 0.539 | 397 / 7311 (5.4%) | 1.261 [1.132, 1.405] | <0.001 |
| Most days or everyday | 47 / 372 (12.6%) | 1.274 [0.927, 1.751] | 0.136 | 80 / 695 (11.5%) | 3.035 [2.376, 3.876] | <0.001 |
| **Staff care for infected/uninfected residents** |  |  |  |  |  |  |
| Not at all | 3112 / 30330 (10.3%) | 1 | NA | 1653 / 43123 (3.8%) | 1 | NA |
| Rarely or sometimes | 5008 / 34797 (14.4%) | 1.114 [1.06, 1.172] | <0.001 | 2758 / 53253 (5.2%) | 1.084 [1.016, 1.157] | 0.015 |
| Often or all the time | 4458 / 25950 (17.2%) | 1.299 [1.233, 1.369] | <0.001 | 2471 / 38870 (6.4%) | 1.204 [1.126, 1.288] | <0.001 |
| Not applicable | 417 / 22083 (1.9%) | 0.308 [0.276, 0.343] | <0.001 | 169 / 28585 (0.6%) | 0.246 [0.209, 0.29] | <0.001 |
| Risk Factor | Proportion with infection (%) | Adjusted OR (95% CI) | p-value | Proportion with infection (%) | Adjusted OR (95% CI) | p-value |
| **Cleaning frequency - communal areas** |  |  |  |  |  |  |
| At least twice per day | 9487 / 83396 (11.4%) | 1 | NA | 5035 / 119764 (4.2%) | 1 | NA |
| Once per day | 3193 / 26511 (12.0%) | 1.053 [1.003, 1.107] | 0.039 | 1802 / 39301 (4.6%) | 1.101 [1.034, 1.172] | 0.003 |
| Other | 315 / 3253 (9.7%) | 0.7 [0.604, 0.81] | <0.001 | 214 / 4766 (4.5%) | 1.213 [1.019, 1.445] | 0.030 |
| **Cleaning frequency - communal touchpoints** |  |  |  |  |  |  |
| At least twice per day | 11472 / 98778 (11.6%) | 1 | NA | 6180 / 144102 (4.3%) | 1 | NA |
| Once per day | 930 / 9648 (9.6%) | 0.85 [0.787, 0.919] | <0.001 | 539 / 12907 (4.2%) | 0.99 [0.897, 1.092] | 0.841 |
| Other | 593 / 4734 (12.5%) | 1.146 [1.025, 1.281] | 0.017 | 332 / 6822 (4.9%) | 1.033 [0.895, 1.192] | 0.655 |
| **Cleaning frequency - staff rooms** |  |  |  |  |  |  |
| At least twice per day | 6186 / 56532 (10.9%) | 1 | NA | 3522 / 81868 (4.3%) | 1 | NA |
| Once per day | 5733 / 48225 (11.9%) | 1.021 [0.977, 1.066] | 0.360 | 3012 / 69493 (4.3%) | 0.905 [0.856, 0.957] | <0.001 |
| Other | 1076 / 8403 (12.8%) | 1.237 [1.144, 1.337] | <0.001 | 517 / 12470 (4.1%) | 0.854 [0.77, 0.946] | 0.002 |
| **Staff use of PPE** |  |  |  |  |  |  |
| All the time | 9696 / 79786 (12.2%) | 1 | NA | 5286 / 115831 (4.6%) | 1 | NA |
| Any contact - all residents | 1132 / 12556 (9.0%) | 0.858 [0.811, 0.907] | <0.001 | 544 / 18129 (3.0%) | 0.924 [0.862, 0.99] | 0.025 |
| Any contact - infected/shielding residents | 124 / 1803 (6.9%) | 1.197 [1.046, 1.37] | 0.009 | 50 / 2481 (2.0%) | 0.886 [0.725, 1.083] | 0.237 |
| Direct care - all residents | 1765 / 16985 (10.4%) | 0.91 [0.85, 0.974] | 0.007 | 1064 / 24526 (4.3%) | 0.824 [0.752, 0.904] | <0.001 |
| Direct care - infected/shielding residents | 278 / 2030 (13.7%) | 0.578 [0.479, 0.699] | <0.001 | 107 / 2864 (3.7%) | 0.512 [0.385, 0.68] | <0.001 |
| **Barrier nursing for infected residents** |  |  |  |  |  |  |
| No | 873 / 34565 (2.5%) | 1 | NA | 521 / 44708 (1.2%) | 1 | NA |
| Yes | 12122 / 78595 (15.4%) | 3.599 [3.337, 3.881] | <0.001 | 6530 / 119123 (5.5%) | 2.598 [2.358, 2.863] | <0.001 |
| **Barrier nursing for all residents** |  |  |  |  |  |  |
| No | 4039 / 49269 (8.2%) | 1 | NA | 2129 / 69668 (3.1%) | 1 | NA |
| Yes | 8956 / 63891 (14.0%) | 1.422 [1.365, 1.482] | <0.001 | 4922 / 94163 (5.2%) | 1.385 [1.312, 1.461] | <0.001 |
| **Inability to isolate a resident (non-compliance)** |  |  |  |  |  |  |
| No | 5993 / 69344 (8.6%) | 1 | NA | 2999 / 98839 (3.0%) | 1 | NA |
| Yes | 7002 / 43816 (16.0%) | 1.328 [1.276, 1.381] | <0.001 | 4052 / 64992 (6.2%) | 1.484 [1.41, 1.563] | <0.001 |
| **Admissions to the LTCF** |  |  |  |  |  |  |
| Risk Factor | Proportion with infection (%) | Adjusted OR (95% CI) | p-value | Proportion with infection (%) | Adjusted OR (95% CI) | p-value |
| Baseline | NA | 1 | NA | NA | 1 | NA |
| Each one unit increase in admissions | 12995 / 113160 (11.5%) | 1.012 [1.01, 1.014] | <0.001 | 7051 / 163831 (4.3%) | 1.004 [1.002, 1.007] | 0.001 |
| **Week of closure to visitors** |  |  |  |  |  |  |
| Baseline (March 1<sup>st</sup>) | NA | 1 | NA | NA | 1 | NA |
| Each additional week since baseline | 12995 / 113160 (11.5%) | 1.02 [1.004, 1.035] | 0.012 | 7051 / 163831 (4.3%) | 1.015 [0.995, 1.034] | 0.143 |

Note: The resident model includes 3311 LTCFs ( $R^2 = 0.27$ ). The staff model includes 3138 LTCFs ( $R^2 = 0.26$ ).

**Table 3.** Risk Factors for large outbreaks in staff or residents of Long Term Care Facilities (LTCFs)

| **Risk Factor** | **Proportion of LTCFs with ≥ 1case** | **Adjusted OR (95% CI)** | **p-value** | **Proportion of LTCFs with a large outbreak** | **Adjusted OR (95% CI)** | **p-value** |
| --- | --- | --- | --- | --- | --- | --- |
| **Social deprivation** |  |  |  |  |  |  |
| All others | 1510 / 2558 (59.0%) | 1 | NA | 231 / 1510 (15.3%) | 1 | NA |
| Most socially deprived quintile | 371 / 571 (65.0%) | 0.983 [0.747, 1.293] | 0.902 | 79 / 371 (21.3%) | 1.322 [0.951, 1.836] | 0.097 |
| **Care Sector** |  |  |  |  |  |  |
| Not for profit | 324 / 499 (64.9%) | 1 | NA | 42 / 324 (13.0%) | 1 | NA |
| For profit | 1557 / 2630 (59.2%) | 1.375 [0.991, 1.909] | 0.057 | 268 / 1557 (17.2%) | 1.647 [1.067, 2.543] | 0.024 |
| **Number of LTCFs in chain** |  |  |  |  |  |  |
| Single | 790 / 1453 (54.4%) | 1 | NA | 120 / 790 (15.2%) | 1 | NA |
| 2 - 9 | 563 / 922 (61.1%) | 0.927 [0.735, 1.17] | 0.522 | 91 / 563 (16.2%) | 0.89 [0.647, 1.224] | 0.473 |
| 10 or more | 528 / 754 (70.0%) | 1.138 [0.86, 1.505] | 0.366 | 99 / 528 (18.8%) | 1.177 [0.842, 1.645] | 0.341 |
| **Ratio of staff to beds** |  |  |  |  |  |  |
| Baseline | NA | 1 | NA | NA | 1 | NA |
| Each one unit increase in staff to bed ratio | 1881 / 3129 (60.1%) | 1.058 [0.878, 1.274] | 0.556 | 310 / 1881 (16.5%) | 1.022 [0.802, 1.302] | 0.862 |
| **Region** |  |  |  |  |  |  |
| London | 136 / 165 (82.4%) | 1 | NA | 8 / 136 (5.9%) | 1 | NA |
| East Midlands | 174 / 297 (58.6%) | 0.28 [0.154, 0.507] | <0.001 | 22 / 174 (12.6%) | 2.548 [1.071, 6.06] | 0.034 |
| East of England | 195 / 346 (56.4%) | 0.275 [0.154, 0.491] | <0.001 | 32 / 195 (16.4%) | 3.287 [1.428, 7.567] | 0.005 |
| North East | 139 / 198 (70.2%) | 0.358 [0.185, 0.692] | 0.002 | 38 / 139 (27.3%) | 5.65 [2.415, 13.217] | <0.001 |
| North West | 282 / 430 (65.6%) | 0.39 [0.218, 0.697] | 0.001 | 43 / 282 (15.2%) | 2.79 [1.232, 6.319] | 0.014 |
| South East | 323 / 554 (58.3%) | 0.289 [0.166, 0.504] | <0.001 | 50 / 323 (15.5%) | 3.092 [1.393, 6.862] | 0.006 |
| South West | 156 / 428 (36.4%) | 0.109 [0.062, 0.193] | <0.001 | 30 / 156 (19.2%) | 4.284 [1.845, 9.946] | 0.001 |
| West Midlands | 261 / 371 (70.4%) | 0.339 [0.188, 0.611] | <0.001 | 50 / 261 (19.2%) | 3.885 [1.738, 8.686] | 0.001 |
| Yorkshire & Humber | 215 / 340 (63.2%) | 0.332 [0.184, 0.6] | <0.001 | 37 / 215 (17.2%) | 3.315 [1.451, 7.575] | 0.004 |
| **Number of beds in care home** |  |  |  |  |  |  |
| Less than 25 | 159 / 503 (31.6%) | 1 | NA | 21 / 159 (13.2%) | 1 | NA |
| 25 to 50 | 959 / 1653 (58.0%) | 1.729 [1.297, 2.305] | <0.001 | 125 / 959 (13.0%) | 0.695 [0.407, 1.187] | 0.183 |
| More than 50 | 763 / 973 (78.4%) | 2.764 [1.967, 3.884] | <0.001 | 164 / 763 (21.5%) | 1.134 [0.658, 1.957] | 0.650 |
| **Quality rating on leadership** |  |  |  |  |  |  |
| Outstanding or good | 1339 / 2292 (58.4%) | 1 | NA | 225 / 1339 (16.8%) | 1 | NA |
| Inadequate or requires improvement | 515 / 793 (64.9%) | 1.169 [0.925, 1.478] | 0.190 | 82 / 515 (15.9%) | 0.826 [0.613, 1.114] | 0.210 |
| No rating | 27 / 44 (61.4%) | 0.791 [0.343, 1.824] | 0.582 | 3 / 27 (11.1%) | 0.568 [0.163, 1.983] | 0.375 |
| **Main type of care provided** |  |  |  |  |  |  |
| Residents aged > 65 years | 1405 / 2406 (58.4%) | 1 | NA | 221 / 1405 (15.7%) | 1 | NA |
| Residents with dementia | 476 / 723 (65.8%) | 1.009 [0.794, 1.282] | 0.942 | 89 / 476 (18.7%) | 0.993 [0.741, 1.332] | 0.965 |
| **Sickness pay** |  |  |  |  |  |  |
| None | 134 / 224 (59.8%) | 1 | NA | 31 / 134 (23.1%) | 1 | NA |
| Statutory | 1415 / 2421 (58.4%) | 0.997 [0.684, 1.452] | 0.987 | 232 / 1415 (16.4%) | 0.593 [0.377, 0.934] | 0.024 |
| Full or more than statutory | 332 / 484 (68.6%) | 1.173 [0.74, 1.858] | 0.497 | 47 / 332 (14.2%) | 0.553 [0.314, 0.973] | 0.040 |
| **Employment of agency nurses or carers** |  |  |  |  |  |  |
| Not employed | 612 / 1372 (44.6%) | 1 | NA | 63 / 612 (10.3%) | 1 | NA |
| Few times a month | 428 / 620 (69.0%) | 1.609 [1.205, 2.148] | 0.001 | 64 / 341 (18.8%) | 1.847 [1.232, 2.77] | 0.003 |
| Few times a week | 341 / 516 (66.1%) | 1.622 [1.234, 2.131] | 0.001 | 69 / 428 (16.1%) | 1.514 [1.023, 2.243] | 0.038 |
| Most days or everyday | 500 / 621 (80.5%) | 2.331 [1.718, 3.163] | <0.001 | 114 / 500 (22.8%) | 2.421 [1.669, 3.511] | <0.001 |
| **Employment of other bank/agency staff** |  |  |  |  |  |  |
| Not employed | 1373 / 2459 (55.8%) | 1 | NA | 212 / 1373 (15.4%) | 1 | NA |
| Few times a month | 182 / 243 (74.9%) | 1.496 [0.991, 2.257] | 0.055 | 38 / 182 (20.9%) | 1.314 [0.863, 2.001] | 0.203 |
| Few times a week | 163 / 236 (69.1%) | 1.134 [0.771, 1.669] | 0.523 | 35 / 163 (21.5%) | 1.391 [0.897, 2.157] | 0.141 |
| Most days or everyday | 163 / 191 (85.3%) | 1.965 [1.17, 3.299] | 0.011 | 25 / 163 (15.3%) | 0.729 [0.446, 1.19] | 0.206 |
| **How often LTCF staff work at other sites** |  |  |  |  |  |  |
| Not at all | 1640 / 2775 (59.1%) | 1 | NA | 268 / 1640 (16.3%) | 1 | NA |
| Few times a month | 140 / 205 (68.3%) | 0.962 [0.635, 1.457] | 0.854 | 26 / 140 (18.6%) | 1.044 [0.647, 1.685] | 0.859 |
| Few times a week | 93 / 137 (67.9%) | 1.146 [0.694, 1.891] | 0.595 | 14 / 93 (15.1%) | 1.044 [0.563, 1.934] | 0.892 |
| Most days everyday | 8 / 12 (66.7%) | 0.802 [0.195, 3.298] | 0.760 | 2 / 8 (25.0%) | 1.771 [0.31, 10.113] | 0.520 |
| **Staff care for uninfected & infected residents** |  |  |  |  |  |  |
| Not at all | 470 / 829 (56.7%) | 1 | NA | 74 / 470 (15.7%) | 1 | NA |
| Rarely or sometimes | 693 / 901 (76.9%) | 1.527 [1.186, 1.965] | 0.001 | 122 / 693 (17.6%) | 0.982 [0.7, 1.379] | 0.917 |
| Often or all the time | 580 / 692 (83.8%) | 2.599 [1.936, 3.488] | <0.001 | 104 / 580 (17.9%) | 0.978 [0.688, 1.391] | 0.902 |
| Not applicable | 138 / 707 (19.5%) | 0.367 [0.278, 0.486] | <0.001 | 10 / 138 (7.2%) | 0.519 [0.25, 1.079] | 0.079 |
| **Cleaning frequency - communal areas** |  |  |  |  |  |  |
| At least twice per day | 1384 / 2331 (59.4%) | 1 | NA | 225 / 1384 (16.3%) | 1 | NA |
| Once per day | 441 / 717 (61.5%) | 1.048 [0.801, 1.369] | 0.734 | 78 / 441 (17.7%) | 1.102 [0.792, 1.532] | 0.564 |
| Other | 56 / 81 (69.1%) | 0.874 [0.397, 1.925] | 0.738 | 7 / 56 (12.5%) | 0.806 [0.299, 2.17] | 0.669 |
| **Cleaning frequency - communal touchpoints** |  |  |  |  |  |  |
| At least twice per day | 1643 / 2741 (59.9%) | 1 | NA | 277 / 1643 (16.9%) | 1 | NA |
| Once per day | 153 / 273 (56.0%) | 0.893 [0.612, 1.302] | 0.556 | 20 / 153 (13.1%) | 0.742 [0.431, 1.276] | 0.280 |
| Other | 85 / 115 (73.9%) | 2.247 [1.108, 4.558] | 0.025 | 13 / 85 (15.3%) | 0.959 [0.448, 2.056] | 0.915 |
| **Cleaning frequency - staff rooms** |  |  |  |  |  |  |
| At least twice per day | 922 / 1573 (58.6%) | 1 | NA | 147 / 922 (15.9%) | 1 | NA |
| Once per day | 802 / 1275 (62.9%) | 0.981 [0.782, 1.232] | 0.871 | 138 / 802 (17.2%) | 1.009 [0.754, 1.35] | 0.952 |
| Other | 157 / 281 (55.9%) | 1.107 [0.757, 1.619] | 0.601 | 25 / 157 (15.9%) | 0.979 [0.586, 1.636] | 0.937 |
| **Staff use of PPE** |  |  |  |  |  |  |
| All the time | 1366 / 2197 (62.2%) | 1 | NA | 228 / 1366 (16.7%) | 1 | NA |
| Direct care all residents | 191 / 375 (50.9%) | 0.961 [0.71, 1.3] | 0.795 | 29 / 191 (15.2%) | 1.03 [0.66, 1.608] | 0.896 |
| Direct care to infected residents | 25 / 47 (53.2%) | 0.681 [0.34, 1.363] | 0.278 | 2 / 25 (8.0%) | 0.483 [0.109, 2.15] | 0.340 |
| Any contact with all residents | 269 / 462 (58.2%) | 0.852 [0.645, 1.125] | 0.258 | 45 / 269 (16.7%) | 0.902 [0.621, 1.31] | 0.589 |
| Any contact with infected residents | 30 / 48 (62.5%) | 0.723 [0.345, 1.518] | 0.392 | 6 / 30 (20.0%) | 1.315 [0.504, 3.43] | 0.576 |
| **Barrier nursing for infected residents** |  |  |  |  |  |  |
| No | 256 / 1083 (23.6%) | 1 | NA | 25 / 256 (9.8%) | 1 | NA |
| Yes | 1625 / 2046 (79.4%) | 5.33 [4.304, 6.601] | <0.001 | 285 / 1625 (17.5%) | 1.288 [0.794, 2.088] | 0.306 |
| **Barrier nursing for all residents** |  |  |  |  |  |  |
| No | 685 / 1387 (49.4%) | 1 | NA | 89 / 685 (13.0%) | 1 | NA |
| Yes | 1196 / 1742 (68.7%) | 1.68 [1.377, 2.049] | <0.001 | 221 / 1196 (18.5%) | 1.437 [1.081, 1.911] | 0.013 |
| **Unable to isolate residents (non-compliance)** |  |  |  |  |  |  |
| No | 1035 / 2052 (50.4%) | 1 | NA | 128 / 1035 (12.4%) | 1 | NA |
| Yes | 846 / 1077 (78.6%) | 1.842 [1.478, 2.296] | <0.001 | 182 / 846 (21.5%) | 1.616 [1.239, 2.108] | <0.001 |
| **Admissions to the LTCF** |  |  |  |  |  |  |
| Baseline | NA | 1 | NA | NA | 1 | NA |
| Each one unit increase in new admissions | 1881 / 3129 (60.1%) | 1.075 [1.05, 1.102] | <0.001 | 310 / 1881 (16.5%) | 1.007 [0.994, 1.02] | 0.292 |
| **Week of closure to visitors<sup>a</sup>** |  |  |  |  |  |  |

<sup>a</sup> Relative to pandemic start

| **Risk Factor** | **Proportion of LTCFs with $\geq 1$ case** | **Adjusted OR (95% CI)** | **p-value** | **Proportion of LTCFs with a large outbreak** | **Adjusted OR (95% CI)** | **p-value** |
| --- | --- | --- | --- | --- | --- | --- |
| Baseline | NA | 1 | NA | NA | 1 | NA |
| Each one unit increase in week | 1881 / 3129 (60.1%) | 0.993 [0.921, 1.071] | 0.854 | 310 / 1881 (16.5%) | 1.058 [0.959, 1.168] | 0.260 |

Note: Cases vs. no cases model is based on 3129 LTCFs, of which 1881 had cases ( $R^2 = 0.39$ ). Large outbreak criteria is LTCFs with more than a third of total number of residents and staff combined testing positive or more than 20 positive combined residents and staff tests. Large vs. small outbreaks model is based on 1881 LTCFs, of which 310 LTCFs were considered to have a "large outbreak". $R^2 = 0.09$ .

### Staffing patterns and practices

Frequent employment of agency nurses or carers (most days or every day versus never) was associated with increased odds infection in residents (aOR: 1·65, 95% CI: 1·56-1·74, p<0·001) and staff (aOR: 1·85, 95% CI: 1·72-1·98, p<0.001), increased odds of ≥1 case in the LTCF (aOR: 2·33, 95% CI: 1·72-3·16, p<0·001) and large outbreaks (aOR: 2·42, 95% CI: 1·67-3·51, p<0·001), Figure 2. Odds of infection were also higher when staff regularly worked across multiple locations (few times per week versus never: aOR: 1·26, 95% CI: 1·13-1·41, p<0·001), Table 2. By comparison with sites that always cohorted staff with infected or uninfected residents, LTCFs in which staff often or always cared for both infected and uninfected residents had higher odds of infection in residents (aOR: 1·30, 95% CI: 1·23-1·37, p<0·001) and staff (aOR: 1·20, 95% CI: 1·13-1·29, p<0·001), and were more likely to have ≥1 infected case (aOR 2·60, 95% CI: 1·94-3·49, p<0.001), but not large outbreaks.

**Figure 2:**
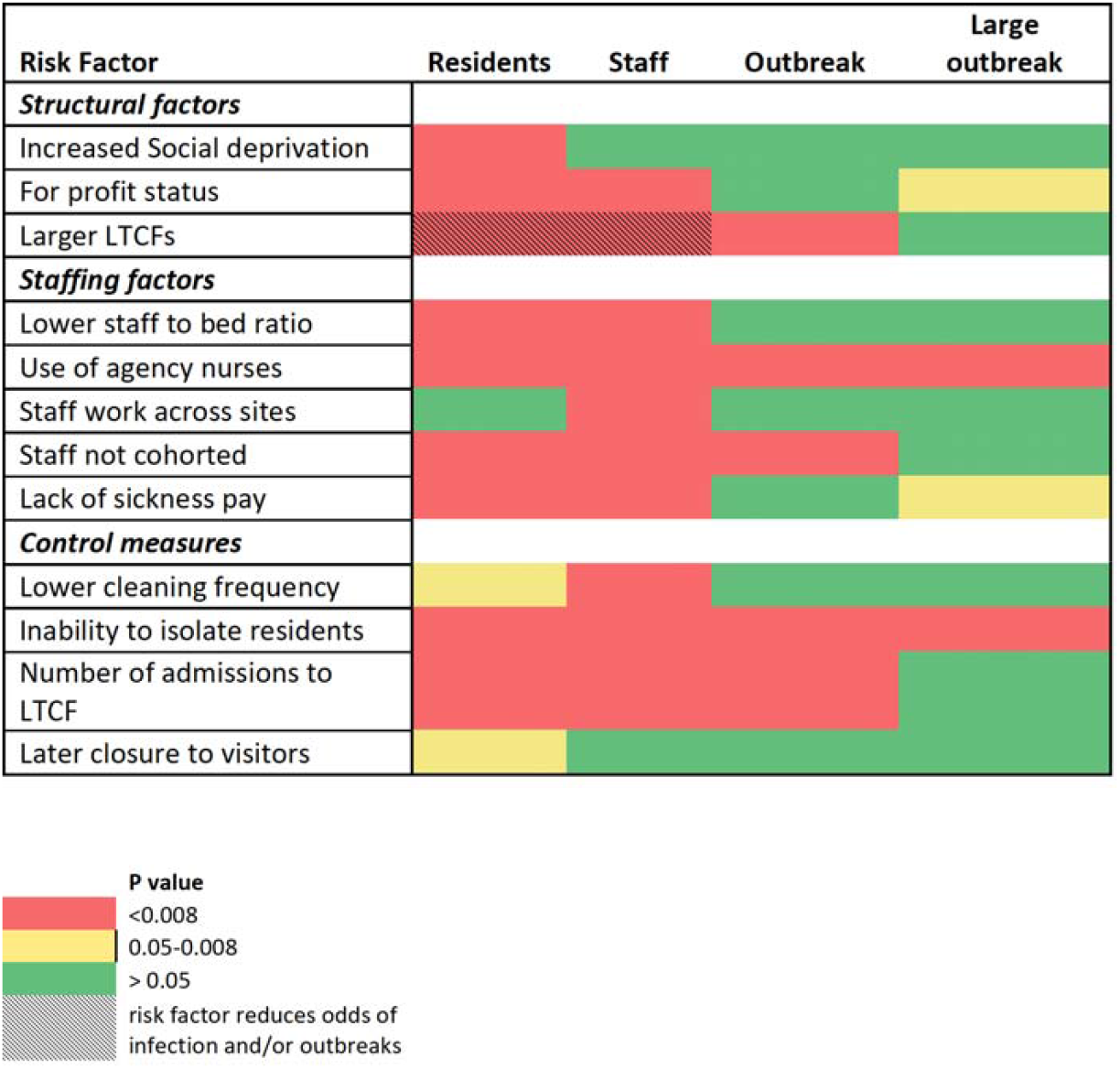
Heat map illustrating main risk factors for infection and outbreaks. The figure above illustrates which risk factors are most strongly associated with infection in residents and staff, outbreaks and large outbreaks. Red denotes strong evidence of an association with the specified risk factor, yellow denotes some evidence of an association, and green denotes no evidence of an association. Each of the columns represents findings from each of the four risk factor analyses. “Residents” is the analysis of risk factors for infection in residents, “Staff” is the analysis of risk factors for infection in staff, “outbreak” is the analysis of risk factors for having at least one COVID-19 case per care home, and “large outbreak” is the analysis of risk factors for large outbreaks rather than small outbreaks.^1^

Protective factors included payment of statutory sickness pay which reduced the odds of infection in residents (aOR: 0·80, 95% CI: 0·75-0·86, p<0.001) and staff (aOR: 0·70, 95% CI: 0·65-0·77, p<0·001), by comparison with LTCFs that did not make this payment, Table 2. Sickness pay also reduced the odds of large outbreaks (OR 0·59, 95% CI: 0·38-0·93, p=0·02), Table 3, but this did not reach statistical significance. Each one unit increase in the staff to bed ratio was associated with reduced odds of infection in residents (OR: 0·82, 95% CI: 0·78-0·87, p<0·001) and staff (0·63, 95% CI: 0·59-0·68, p<0·001).

### Use of disease control measures

The inability to isolate residents due to non-compliance, for example due to dementia, was associated with increased odds of infection in residents (aOR: 1·33, 95% CI: 1·28-1·38 p<0·001), staff (aOR: 1·48, 95% CI: 1·41-1·56, p<0·001), large outbreaks (aOR 1·62, 95% CI: 1·24-2·11, p<0·001), and ≥1 case (aOR: 1·84, 95% CI: 1·48-2·30, p<0·001), Figure 2. LTCFs which reported cleaning communal areas less frequently than twice per day had higher odds of infection in staff (aOR; 1·10, 95% CI: 1·03-1·17, p=0·003), Table 2, with some evidence of an effect in residents (aOR: 1·05, 95% CI: 1·00-1·10, p=0·04). Cleaning frequency in other areas and PPE use were not associated with infection or outbreaks. The use of barrier nursing was strongly associated with higher odds of infection and ≥1 case, and there was moderate evidence of an association with large outbreaks (aOR 1·44, 95% CI: 1·08-1·91, p=0·01), see discussion for potential explanations. The number of admissions to the facility was associated with increased odds of infection in residents (aOR: 1·01, 95% CI: 1·01-1·01, p<0·001), staff (aOR: 1·00, 95% CI: 1·00-1·00, p<0·001), and the odds of ≥1 case in the LTCF (aOR 1·08, 95% CI: 1·05-1·10, p<0·001), Table 3. There was moderate evidence that later closure to visitors was a risk factor for infection in residents (aOR 1·02, 95% CI: 1·00-1·04, p=0·01).

Analysis of risk factors for infection using the survey-test dataset found an association between increasing age (per year) and the odds of infection in residents (aOR: 1·01, 95% CI: 1·01-1·03, p<0·001) but not staff. Similar risk factors for infection were identified when the analysis was repeated using test results as the outcome, eTable2, and in sensitivity analysis using multiple imputation to account for missing survey data, eTables3-5.

## Discussion

This national survey demonstrates the widespread but variable impact of COVID-19 on LTCFs in England, as almost half of LTCFs did not report any cases. This emphasises the need for effective infection prevention and control strategies because many facilities remain vulnerable to infection. Our findings highlight the key role of staff, compliance with control measures, and new admissions in transmission of infection, and are consistent with the long-recognised drivers of infection in LTCFs such as overcrowding, contact frequency, and staffing ratios.^20–23^ We identify specific strategies that could be deployed immediately to reduce the risk of COVID-19 in LTCFs, including provision of financial support to staff so they are incentivised to test and self-isolate when unwell, reduced use of agency staff, improved staffing ratios, and widespread adoption of disease control measures such as cohorting and isolation.

Likelihood of infection and outbreaks was reduced in LTCFs which cohorted staff with either infected or uninfected residents, in common with a recent survey of 132 LTCFs in South West France.^24^ Facilities that provided staff sickness pay and did not employ agency staff had fewer cases of infection. Infections in staff were also more likely in LTCFs in which staff regularly worked across multiple settings, which is consistent with an English study which found three-fold higher odds of infection in staff who worked across multiple locations compared to those who worked at one site.^25^ Taken together this provides compelling evidence that staff play a key role in transmitting infection to each other and to residents, although it is difficult to discern a causal relationship between employment of agency staff and initiation of outbreaks without dates of employment and infection. Likelihood of infection in staff and residents was also driven by new admissions to the LTCFs, highlighting the potential risks of importation of infection by new or returning residents.

Almost half of LTCFs in this study did not report any infections and similar findings from regional studies in North America and Europe suggest that many staff and residents remain vulnerable to COVID-19^11,24,26,27^. Prevention of future infections requires coordinated investment to recompense staff who are required to shield or self-isolate so that they do not take on multiple jobs, and investment to minimise reliance on agency staff and reduce the number of individuals who work across multiple locations. In England, the Government has recently established the Infection Control Fund which provides regions with financial support to cover the costs of implementing these measures.^28^ Given the critical role of staff in transmission of infection, our finding of high prevalence of asymptomatic infection in this group,^29^ emphasise the importance of regular staff testing to facilitate early detection of infection and prevent outbreaks. Strategies are also required to improve the implementation of disease control measures such as isolation. The association between admissions and infection points to the importance of testing and isolating residents on admission to the LTCF.

## Limitations

In common with all cross-sectional studies, our findings may be subject to confounding and reverse causality and this is particularly problematic for measures which may be initiated in response to infection, such as barrier nursing or employment of agency staff. Variability in access to testing capacity and earlier onset of the pandemic in London are likely explanations for the regional differences in prevalence that were observed in staff and residents. Managers of LTCFs were asked to report on behalf of their staff and residents, and it is possible that social desirability bias affected survey responses. Confidence in our results is increased by evaluation of risk factors for infection and outbreaks, which allowed us to investigate factors associated with both ingress and spread of infection, and the fact that similar results were obtained in sensitivity analyses. The national scale of our study, and comparability of survey responders and non-responders supports our assertion that findings can be generalized to all LTCFs in England.

## Conclusions

This study documented the substantial burden of SARS-CoV-2 across LTCFs, but also identified half of LTCFs without outbreaks which remain vulnerable to infection. Strategies that could be implemented immediately to reduce the impact of SARS-CoV-2 in LTCFs include: provision of financial support to the LTCF workforce to incentivise testing and self-isolation when sick, investment to reduce reliance on agency staff, and a focus on implementing disease control measures such as cohorting and isolation. Widespread, rapid adoption of these measures could support efforts to protect this vulnerable sector of society from future waves of infection.

## Data Availability

De-identified, LTCF-level data collected in the survey data and the associated questionnaire will be deposited in the ONS Secure Research Service (SRS) at the time of publication in a peer reviewed journal, or shortly afterwards, for use by accredited researchers. The study protocol is also available on the UCL website (https://doi.org/10.5522/04/12993506.v1). Accredited researchers can access the survey via the SRS following submission of a project proposal which sets out the intended use of the dataset and the value of the project. Further details about applying to use the SRS are available from: https://www.ons.gov.uk/aboutus/whatwedo/statistics/requestingstatistics/approvedresearcherscheme

## Acknowledgements

The authors would like to acknowledge Rachel Williams and Anna Quigley from Ipsos MORI who led the survey fieldwork and Julian Sandler from the Office for National Statistics who supported construction of the Tables. This report is independent research funded by the Department of Health and Social Care (COVID-19 Surveillance studies). The views expressed in this publication are those of the authors and not necessarily those of the NHS, or the Department of Health and Social Care.

## Author contributions

LS, AH, and SH designed the survey. OA and LW designed and piloted the questionnaire. OA, ST, DB and LS designed the statistical analysis plan and DB, ST, KS, JS undertook the statistical analysis. AD managed the project. GH extracted the dataset and undertook the data linkage under instructions from the research team. MK reviewed the literature. LS wrote the first draft of the manuscript. All authors revised and edited the manuscript.

## Footnotes

1 Small outbreak= at least one case; large outbreak defined above

