## Supplementary Tables and Methods for "Risk factors associated with SARS-CoV-2 infection and outbreaks in Long Term Care Facilities in England: a national survey"

**Online-only supplement**

**eMethods: Linkage of individual-level test results to the survey**

**eTable1: Laboratory-confirmed SARS-CoV-2 prevalence by age-group in residents and staff who participated in the whole care home testing programme (30^th^ April -13 June 2020). A) Weighted estimates and B) raw data**

**eTable2: Multivariable analysis of risk factors for laboratory confirmed SARS-CoV-2 infection in staff and residents based on the linked test-survey dataset**

**eTable 3. Multivariable analysis of risk factors for infection in residents and staff (using survey data as the outcome) including multiple imputation**

**eTable4: Multivariable analysis of risk factors for infection in residents and staff (using the linked test-survey dataset) with multiple imputation**

**eTable5: Multivariable analysis of risk factors for large or prolonged outbreaks in staff or residents after multiple imputation**

**eMethods: Linkage of individual-level test results to survey**

Test results were obtained through the Pillar 2 testing programme, which provides community testing for individuals in England and includes testing for residents and staff of Long Term Care Facilities (LTCFs). Before the national testing programme in LTCFs was launched in April 30^th^, national policy was to only test symptomatic residents and staff, and testing was limited to a maximum of 5 samples per facility. From 30 April onwards, all LTCFs were expected to take part in the national testing programme. We therefore aimed to use test results from residents and staff to estimate the point prevalence of SARS-CoV-2, by identifying samples collected when each LTCF participated in the whole care home testing programme. As staff and residents were only expected to submit one sample per person in this programme, this reduced the risk of including duplicate samples from the same individual (as it was not possible to link test results to personal identifiers).

We took the following steps to identify samples from LTCF residents and staff and to link survey responses to individual-level testing data, see Figure 1 in the manuscript.

1. Pillar 2 test results were attributed to staff or residents of LTCFs if they met any of the following criteria:

- Care Quality Commission (CQC) identifier (linking the sample to a specific LTCF)
- Sample labelled as being obtained from a LTCF resident or staff member
- Test site recorded as LTCF

2. Next samples were linked to LTCFs in the sample frame that was used for the survey, to restrict the analysis to LTCFs that primarily provided care to residents aged > 65 years, or to individuals with dementia.
3. Date restrictions were applied to exclude samples obtained outside the whole care home testing period
4. LTCFs which only had tests from symptomatic cases (or had < 10 tests in which symptomatic/asymptomatic status was not recorded) were excluded, because this implied that the LTCF had not participated in the whole care home testing programme
5. The date that the LTCF participated in whole care home testing was available from the survey for one-third of LTCFs. For the remainder, an algorithm was applied to identify the “peak” period of testing, since this was likely to represent when each LTCF had participated in the whole care home testing programme. Test results obtained either side of this peak were included until there was a day with no tests (excluding weekends when testing is rarely undertaken in LTCFs). The algorithm’s performance was compared to actual dates of testing for the subset of LTCFs for which this information was available, and in 96.5% of these LTCFs the estimated date of peak testing was within 3 days of the actual date of testing that was reported in the survey. This reflects that testing was usually spread across several days in most LTCFs, due to the requirement to test all staff and residents, and that the LTCF may have reported any date between the first and last day of testing in the survey.
6. Finally we applied rules to remove LTCFs which had substantially more tests than the total number of staff and residents recorded in the survey. LTCFs were removed if the number of resident tests was at least 10 more than the number of residents recorded in the survey and/or the number of staff tests was at least 5 more than the number of staff.

**eTable 1: Laboratory-confirmed SARS-CoV-2 prevalence by age-group in residents and staff who participated in the whole care home testing programme (30^th^ April -13 June 2020). A) Weighted estimates and B) raw data**

A) weighted estimates of prevalence

| **Age-group** | **Proportion positive tests / total tests (95% CI)** | **Proportion asymptomatic positive tests / all positive tests (95% CI)** | **Proportion symptomatic positive tests / all positive tests (95% CI)** |
| --- | --- | --- | --- |
| **RESIDENTS** | | | |
| 0-25 | - | - | - |
| 26-35 | - | - | - |
| 36-45 | - | - | - |
| 46-55 | - | - | - |
| 56-65 | 0.012 [0.009, 0.015] | 0.300 [0.298, 0.302] | 0.000 [0.000, 0.000] |
| 66-75 | 0.024 [0.021, 0.028] | 0.733 [0.724, 0.742] | 0.108 [0.100, 0.115] |
| 76-85 | 0.028 [0.025, 0.032] | 0.795 [0.788, 0.803] | 0.083 [0.077, 0.088] |
| 86-95 | 0.028 [0.025, 0.032] | 0.796 [0.787, 0.804] | 0.078 [0.073, 0.084] |
| 96-105 | 0.026 [0.022, 0.030] | 0.763 [0.755, 0.772] | 0.093 [0.087, 0.099] |
| 106+ | 0.034 [-0.001, 0.068] | 0.030 [0.008, 0.052] | 0.030 [0.008, 0.052] |
| **STAFF** | | | |
| 0-25 | 0.007 [0.004, 0.010] | 0.899 [0.893, 0.905] | 0.050 [0.044, 0.056] |
| 26-35 | 0.005 [0.003, 0.008] | 0.871 [0.865, 0.876] | 0.034 [0.029, 0.040] |
| 36-45 | 0.006 [0.004, 0.009] | 0.827 [0.820, 0.835] | 0.096 [0.089, 0.104] |
| 46-55 | 0.006 [0.003, 0.009] | 0.971 [0.968, 0.975] | 0.017 [0.014, 0.020] |
| 56-65 | 0.006 [0.004, 0.009] | 0.872 [0.865, 0.879] | 0.062 [0.055, 0.068] |
| 66-75 | 0.005 [0.003, 0.006] | 0.476 [0.476, 0.476] | 0.000 [0.000, 0.000] |
| 76-85 | 0.006 [0.005, 0.008] | 0.118 [0.118, 0.118] | 0.000 [0.000, 0.000] |
| 86-95 | - | - | - |
| 96-105 | - | - | - |
| 106+ | - | - | - |

B) Raw test data

| **Age-group** | **Number of positive tests/Number of tests** | **Number of asymptomatic positive tests / Number of positive tests** | **Number of symptomatic positive tests / Number of positive tests** |
| --- | --- | --- | --- |
| **RESIDENTS** | | | |
| 0-25 | - | - | - |
| 26-35 | - | - | - |
| 36-45 | - | - | - |
| 46-55 | - | - | - |
| 56-65 | 2 / 262 | 2/2 | 0 / 2 |
| 66-75 | 154 / 6164 | 121 / 154 | 14 / 154 |
| 76-85 | 499 / 18507 | 386 / 499 | 41 / 499 |
| 86-95 | 813 / 29169 | 629 / 813 | 61 / 813 |
| 96-105 | 158 / 6460 | 119 / 158 | 16 / 158 |
| 106+ | 2/50 | 0 / 2 | 1/2 |
| **STAFF** | | | |
| 0-25 | 45 / 6875 | 42 / 45 | 3 / 45 |
| 26-35 | 60 / 10164 | 58 / 60 | 2 / 60 |
| 36-45 | 66 / 10419 | 62 / 66 | 4 / 66 |
| 46-55 | 80 / 13021 | 80 / 80 | 0 / 80 |
| 56-65 | 67 / 10812 | 65 / 67 | 2 / 67 |
| 66-75 | 3 / 1579 | 3 / 3 | 0 / 3 |
| 76-85 | 2 / 118 | 2 / 2 | 0 / 2 |
| 86-95 | - | - | - |
| 96-105 | - | - | - |
| 106+ | - | - | - |

**eTable2: Multivariable analysis of risk factors for laboratory confirmed SARS-CoV-2 infection in staff and residents based on the linked test-survey dataset**

| **Risk Factor** | **Residents** | | | | **Staff** | | | |
| --- | --- | --- | --- | --- | --- | --- | --- | --- |
|  | **Number per category (%)** | **Proportion with infection** | **Adjusted OR (95% CI)** | **p-value** | **Number per category (%)** | **Proportion with infection (%)** | **Adjusted OR (95% CI)** | **p-value** |
| **Age^[1]^** |  |  |  |  |  |  |  |  |
| Baseline age-group^[2]^ |  |  | 1 |  |  |  | 1 |  |
| Each one year increase in age | 40613 (100%) | 1022 / 40613 (2.5%) | 1.016 [1.007, 1.026] | 0.001 | 34874 (100%) | 226 / 34874 (0.6%) | 0.997 [0.987, 1.007] | 0.581 |
| **Gender^a^** |  |  |  |  |  |  |  |  |
| Female | 29141 (71.8%) | 736 / 29141 (2.5%) | 1 |  | 29082 (83.4%) | 190 / 29082 (0.7%) | 1 |  |
| Male | 11472 (28.2%) | 286 / 11472 (2.5%) | 1.027 [0.878, 1.201] | 0.743 | 5792 (16.6%) | 36 / 5792 (0.6%) | 0.883 [0.609, 1.28] | 0.510 |
| **Social deprivation** |  |  |  |  |  |  |  |  |
| All others | 1364 (85.1%) | 837 / 34317 (2.4%) | 1 |  | 962 (89.7%) | 201 / 31022 (0.6%) | 1 |  |
| Most socially deprived quintile | 239 (14.9%) | 185 / 6296 (2.9%) | 1.068 [0.733, 1.556] | 0.731 | 110 (10.3%) | 25 / 3852 (0.6%) | 0.796 [0.384, 1.65] | 0.540 |
| **Care sector** |  |  |  |  |  |  |  |  |
| Not for profit | 242 (15.1%) | 189 / 6875 (2.7%) | 1 |  | 148 (13.8%) | 32 / 5997 (0.5%) | 1 |  |
| For profit | 1361 (84.9%) | 833 / 33738 (2.5%) | 1.232 [0.788, 1.926] | 0.360 | 924 (86.2%) | 194 / 28877 (0.7%) | 1.047 [0.491, 2.233] | 0.905 |
| **Number of LTCFs in group** |  |  |  |  |  |  |  |  |
| Single | 769 (48%) | 371 / 17255 (2.2%) | 1 |  | 563 (52.5%) | 113 / 16808 (0.7%) | 1 |  |
| 2-9 | 432 (26.9%) | 245 / 10984 (2.2%) | 0.953 [0.678, 1.339] | 0.782 | 204 (19.0%) | 63 / 10280 (0.6%) | 0.753 [0.39, 1.453] | 0.398 |
| 10 or more | 402 (25.1%) | 406 / 12374 (3.3%) | 1.439 [1.001, 2.067] | 0.049 | 305 (28.5%) | 50 / 7786 (0.6%) | 0.733 [0.431, 1.246] | 0.251 |
| **Staff to bed ratio** |  |  |  |  |  |  |  |  |
| Baseline |  |  | 1 |  |  |  | 1 |  |
| Each one unit increase in staff-bed ratio | 1603 (100%) | 1022 / 40613 (2.5%) | 0.746 [0.493, 1.128] | 0.165 | 1072 (100%) | 226 / 34874 (0.6%) | 0.963 [0.57, 1.625] | 0.886 |
| **Region** |  |  |  |  |  |  |  |  |
| London | 87 (5.4%) | 56 / 2455 (2.3%) | 1 |  | 71 (6.6%) | 7 / 2235 (0.3%) | 1 |  |
| East Midlands | 166 (10.4%) | 80 / 3979 (2.0%) | 0.966 [0.482, 1.938] | 0.923 | 114 (10.6%) | 21 / 3612 (0.6%) | 2.465 [0.742, 8.191] | 0.141 |
| East of England | 194 (12.1%) | 135 / 5195 (2.6%) | 1.132 [0.577, 2.221] | 0.719 | 176 (16.4%) | 38 / 5563 (0.7%) | 2.427 [0.764, 7.704] | 0.133 |
| North East | 71 (4.4%) | 102 / 1822 (5.6%) | 2.608 [1.218, 5.588] | 0.014 | 10 (0.9%) | 2 / 274 (0.7%) | 1.426 [0.119, 17.097] | 0.780 |
| North West | 173 (10.8%) | 142 / 4759 (3.0%) | 1.53 [0.788, 2.969] | 0.209 | 26 (2.4%) | 11 / 889 (1.2%) | 5.184 [1.224, 21.958] | 0.025 |
| South East | 350 (21.8%) | 196 / 8216 (2.4%) | 1.137 [0.612, 2.114] | 0.685 | 304 (28.4%) | 75 / 10095 (0.7%) | 2.941 [1.002, 8.633] | 0.050 |
| South West | 258 (16.1%) | 49 / 6388 (0.8%) | 0.464 [0.228, 0.945] | 0.034 | 216 (20.1%) | 21 / 7140 (0.3%) | 1.323 [0.402, 4.351] | 0.645 |
| West Midlands | 174 (10.9%) | 111 / 4596 (2.4%) | 0.982 [0.496, 1.944] | 0.959 | 118 (11.0%) | 25 / 4015 (0.6%) | 2.197 [0.659, 7.327] | 0.200 |
| Yorkshire & Humber | 130 (8.1%) | 151 / 3203 (4.7%) | 2.446 [1.231, 4.859] | 0.011 | 37 (3.5%) | 26 / 1051 (2.5%) | 12.166 [3.196, 46.307] | <0.001 |
| **Number of care home beds** |  |  |  |  |  |  |  |  |
| Less than 25 | 247 (15.4%) | 34 / 3129 (1.1%) | 1 |  | 200 (18.7%) | 10 / 3473 (0.3%) | 1 |  |
| 25-50 | 818 (51%) | 459 / 18425 (2.5%) | 1.108 [0.629, 1.952] | 0.722 | 547 (51.0%) | 82 / 16129 (0.5%) | 1.363 [0.569, 3.266] | 0.487 |
| More than 50 | 538 (33.6%) | 529 / 19059 (2.8%) | 1.063 [0.59, 1.915] | 0.839 | 325 (30.3%) | 134 / 15272 (0.9%) | 1.817 [0.736, 4.482] | 0.195 |
| **Quality rating on leadership** |  |  |  |  |  |  |  |  |
| Outstanding or good | 1163 (72.6%) | 730 / 30334 (2.4%) | 1 |  | 776 (72.4%) | 161 / 25847 (0.6%) | 1 |  |
| Requires improvement / inadequate | 418 (26.1%) | 281 / 9840 (2.9%) | 0.991 [0.723, 1.36] | 0.956 | 277 (25.8%) | 61 / 8491 (0.7%) | 1.021 [0.61, 1.707] | 0.937 |
| No rating | 22 (1.4%) | 11 / 439 (2.5%) | 0.765 [0.211, 2.777] | 0.684 | 19 (1.8%) | 4 / 536 (0.7%) | 1.12 [0.224, 5.605] | 0.890 |
| **Main type of care provided** |  |  |  |  |  |  |  |  |
| Residents aged > 65 years | 1255 (78.3%) | 778 / 31028 (2.5%) | 1 |  | 832 (77.6%) | 165 / 26372 (0.6%) | 1 |  |
| Residents with dementia | 348 (21.7%) | 244 / 9585 (2.5%) | 0.967 [0.699, 1.338] | 0.840 | 240 (22.4%) | 61 / 8502 (0.7%) | 0.985 [0.582, 1.669] | 0.956 |
| **Sickness pay** |  |  |  |  |  |  |  |  |
| None | 115 (7.2%) | 119 / 3106 (3.8%) | 1 |  | 81 (7.6%) | 16 / 2638 (0.6%) | 1 |  |
| Statutory | 1247 (77.8%) | 730 / 30815 (2.4%) | 0.683 [0.414, 1.124] | 0.134 | 833 (77.7%) | 181 / 26138 (0.7%) | 1.333 [0.546, 3.259] | 0.528 |
| Full or more than statutory | 241 (15%) | 173 / 6692 (2.6%) | 0.668 [0.367, 1.216] | 0.187 | 158 (14.7%) | 29 / 6098 (0.5%) | 0.805 [0.272, 2.383] | 0.695 |
| **Employment of bank/agency nurses or carers** |  |  |  |  |  |  |  |  |
| Not employed | 717 (44.7%) | 292 / 16803 (1.7%) | 1 |  | 488 (45.5%) | 69 / 14398 (0.5%) | 1 |  |
| Employed | 886 (55.3%) | 730 / 23810 (3.1%) | 1.332 [0.979, 1.812] | 0.068 | 584 (54.5%) | 157 / 20476 (0.8%) | 1.602 [0.962, 2.668] | 0.070 |
| **Employment of other agency/bank staff** |  |  |  |  |  |  |  |  |
| Not employed | 1248 (77.9%) | 717 / 31140 (2.3%) | 1 |  | 837 (78.1%) | 160 / 26746 (0.6%) | 1 |  |
| Employed | 355 (22.1%) | 305 / 9473 (3.2%) | 1.309 [0.953, 1.798] | 0.097 | 235 (21.9%) | 66 / 8128 (0.8%) | 1.002 [0.589, 1.705] | 0.993 |
| **How often LTCF staff work at other locations** |  |  |  |  |  |  |  |  |
| Not at all | 1425 (88.9%) | 905 / 35988 (2.5%) | 1 |  | 938 (87.5%) | 186 / 30471 (0.6%) | 1 |  |
| Rarely/sometimes/often | 178 (11.1%) | 117 / 4625 (2.5%) | 1.077 [0.706, 1.643] | 0.730 | 134 (12.5%) | 40 / 4403 (0.9%) | 1.406 [0.759, 2.608] | 0.279 |
| **Staff care for uninfected & infected residents** |  |  |  |  |  |  |  |  |
| Not at all | 446 (27.8%) | 234 / 11525 (2.0%) | 1 |  | 304 (28.4%) | 73 / 9803 (0.7%) | 1 |  |
| Rarely or sometimes | 458 (28.6%) | 363 / 12817 (2.8%) | 1.272 [0.897, 1.805] | 0.177 | 300 (28.0%) | 60 / 10820 (0.6%) | 0.825 [0.464, 1.467] | 0.513 |
| Often or all the time | 343 (21.4%) | 413 / 8737 (4.7%) | 2.091 [1.451, 3.014] | <0.001 | 204 (19.0%) | 81 / 6572 (1.2%) | 1.352 [0.739, 2.472] | 0.328 |
| Not applicable | 356 (22.2%) | 12 / 7534 (0.2%) | 0.189 [0.091, 0.393] | <0.001 | 264 (24.6%) | 12 / 7679 (0.2%) | 0.395 [0.169, 0.925] | 0.032 |
| **Cleaning frequency - communal areas** |  |  |  |  |  |  |  |  |
| Twice a day | 1188 (74.1%) | 703 / 29965 (2.3%) | 1 |  | 802 (74.8%) | 160 / 25368 (0.6%) | 1 |  |
| Other or once per day | 415 (25.9%) | 319 / 10648 (3.0%) | 1.213 [0.856, 1.718] | 0.277 | 270 (25.2%) | 66 / 9506 (0.7%) | 0.738 [0.406, 1.341] | 0.319 |
| **Cleaning frequency- communal touchpoints** |  |  |  |  |  |  |  |  |
| Twice per day | 1398 (87.2%) | 881 / 35130 (2.5%) | 1 |  | 948 (88.4%) | 196 / 30910 (0.6%) | 1 |  |
| Other or once per day | 205 (12.8%) | 141 / 5483 (2.6%) | 0.974 [0.631, 1.503] | 0.905 | 124 (11.6%) | 30 / 3964 (0.8%) | 1.268 [0.58, 2.773] | 0.552 |
| **Cleaning frequency – staff rooms** |  |  |  |  |  |  |  |  |
| Twice per day | 821 (51.2%) | 503 / 20940 (2.4%) | 1 |  | 564 (52.6%) | 111 / 18014 (0.6%) | 1 |  |
| Other or once per day | 782 (48.8%) | 519 / 19673 (2.6%) | 0.892 [0.661, 1.203] | 0.453 | 508 (47.4%) | 115 / 16860 (0.7%) | 0.872 [0.53, 1.434] | 0.589 |
| **Staff use of PPE** |  |  |  |  |  |  |  |  |
| All the time | 1113 (69.4%) | 768 / 28345 (2.7%) | 1 |  | 734 (68.5%) | 164 / 23695 (0.7%) | 1 |  |
| Any contact with residents | 262 (16.3%) | 118 / 5704 (2.1%) | 0.655 [0.444, 0.968] | 0.034 | 147 (13.7%) | 30 / 5042 (0.6%) | 0.815 [0.396, 1.678] | 0.579 |
| Direct contact with residents | 228 (14.2%) | 136 / 6564 (2.1%) | 0.88 [0.575, 1.348] | 0.558 | 191 (17.8%) | 32 / 6137 (0.5%) | 0.738 [0.399, 1.365] | 0.332 |
| **Use of barrier nursing for infected residents** |  |  |  |  |  |  |  |  |
| No | 559 (34.9%) | 76 / 12334 (0.6%) | 1 |  | 398 (37.1%) | 41 / 11490 (0.4%) | 1 |  |
| Yes | 1044 (65.1%) | 946 / 28279 (3.3%) | 3.981 [2.565, 6.178] | <0.001 | 674 (62.9%) | 185 / 23384 (0.8%) | 1.655 [0.881, 3.109] | 0.117 |
| **Use of barrier nursing for all residents** |  |  |  |  |  |  |  |  |
| No | 712 (44.4%) | 336 / 17943 (1.9%) | 1 |  | 483 (45.1%) | 98 / 15740 (0.6%) | 1 |  |
| Yes | 891 (55.6%) | 686 / 22670 (3.0%) | 1.411 [1.062, 1.874] | 0.017 | 589 (54.9%) | 128 / 19134 (0.7%) | 0.731 [0.465, 1.15] | 0.176 |
| **Inability to isolate a resident due to non-compliance** |  |  |  |  |  |  |  |  |
| No | 1058 (66%) | 475 / 25771 (1.8%) | 1 |  | 733 (68.4%) | 111 / 22602 (0.5%) | 1 |  |
| Yes | 545 (34%) | 547 / 14842 (3.7%) | 1.36 [1.026, 1.802] | 0.032 | 339 (31.6%) | 115 / 12272 (0.9%) | 1.626 [1.021, 2.59] | 0.041 |
| **Admissions to the LTCF** |  |  |  |  |  |  |  |  |
| Baseline |  |  | 1 |  |  |  | 1 |  |
| Each one unit increase in admissions | 1603 (100%) | 1022 / 40613 (2.5%) | 1.016 [1, 1.032] | 0.055 | 1072 (100%) | 226 / 34874 (0.6%) | 1.015 [0.991, 1.04] | 0.227 |
| **Week of closure to visitors** |  |  |  |  |  |  |  |  |
| Baseline (March 1^st^) |  |  | 1 |  |  |  | 1 |  |
| Number of weeks since baseline | 1603 (100%) | 1022 / 40613 (2.5%) | 0.906 [0.807, 1.017] | 0.095 | 1072 (100%) | 226 / 34874 (0.6%) | 0.986 [0.82, 1.185] | 0.879 |

Note: The resident model is based on 40613 residents in 1603 LTCFs (Intercept = -7.30, SD = 1.57; Marginal R² = 0.06; Conditional R² = 0.12). The staff model is based on 34874 staff in 1072 LTCFs (Intercept = -7.93, SD = 1.70; Marginal R² = 0.01; Conditional R² = 0.05).

**eTable 3. Multivariable analysis of risk factors for infection in residents and staff (using survey data as the outcome) including multiple imputation**

| **Risk Factor** |  | **Residents** |  |  | **Staff** |  |  |
| --- | --- | --- | --- | --- | --- | --- | --- |
|  | **Number LTCFs (%)** | **Prevalence of infection** | **Adjusted OR (95% CI)** | **p-value** | **Prevalence of infection** | **Adjusted OR (95% CI)** | **p-value** |
| **Social deprivation** |  |  |  |  |  |  |  |
| All others | 3505 (82.7%) | 13231 / 114350 (11.6%) | 1 |  | 7433 / 173977 (4.3%) | 1 |  |
| Most socially deprived quintile | 733 (17.3%) | 3752 / 26030 (14.4%) | 1.117 [1.068, 1.167] | <0.001 | 1831 / 38387 (4.8%) | 0.886 [0.836, 0.939] | <0.001 |
| **Care sector** |  |  |  |  |  |  |  |
| Not for profit | 714 (16.8%) | 3059 / 25371 (12.1%) | 1 |  | 1749 / 44342 (3.9%) | 1 |  |
| For profit | 3524 (83.2%) | 13924 / 115009 (12.1%) | 1.154 [1.096, 1.216] | <0.001 | 7515 / 168022 (4.5%) | 1.153 [1.08, 1.23] | <0.001 |
| **Number of LTCFs in chain** |  |  |  |  |  |  |  |
| Single provider | 2054 (48.5%) | 6193 / 59372 (10.4%) | 1 |  | 3321 / 89373 (3.7%) | 1 |  |
| 2-9 LTCFs | 1227 (29%) | 4951 / 40822 (12.1%) | 0.988 [0.947, 1.031] | 0.587 | 2894 / 63119 (4.6%) | 1.035 [0.981, 1.092] | 0.206 |
| 10+ LTCFs | 957 (22.6%) | 5839 / 40186 (14.5%) | 1.097 [1.05, 1.147] | <0.001 | 3049 / 59872 (5.1%) | 1.071 [1.012, 1.133] | 0.018 |
| **Staff to bed ratio** |  |  |  |  |  |  |  |
| Baseline |  |  | 1 |  |  | 1 |  |
| Each one unit increase in staff : bed ratio | 4238 (100%) | 16983 / 140380 (12.1%) | 0.856 [0.819, 0.894] | <0.001 | 9264 / 212364 (4.4%) | 0.694 [0.656, 0.735] | <0.001 |
| **Region** |  |  |  |  |  |  |  |
| London | 229 (5.4%) | 1270 / 8505 (14.9%) | 1 |  | 396 / 13461 (2.9%) | 1 |  |
| East Midlands | 406 (9.6%) | 1303 / 12706 (10.3%) | 0.939 [0.861, 1.023] | 0.150 | 804 / 18672 (4.3%) | 1.97 [1.739, 2.233] | <0.001 |
| East of England | 470 (11.1%) | 1712 / 16621 (10.3%) | 0.921 [0.849, 0.999] | 0.048 | 893 / 23811 (3.8%) | 1.673 [1.48, 1.891] | <0.001 |
| North East | 248 (5.9%) | 1470 / 9134 (16.1%) | 1.175 [1.076, 1.282] | <0.001 | 1032 / 12763 (8.1%) | 3.248 [2.868, 3.677] | <0.001 |
| North West | 596 (14.1%) | 2662 / 19723 (13.5%) | 0.966 [0.895, 1.044] | 0.382 | 1406 / 28575 (4.9%) | 1.851 [1.647, 2.081] | <0.001 |
| South East | 786 (18.5%) | 2668 / 24700 (10.8%) | 0.911 [0.846, 0.982] | 0.015 | 1558 / 38620 (4.0%) | 1.681 [1.499, 1.884] | <0.001 |
| South West | 589 (13.9%) | 1537 / 17831 (8.6%) | 0.884 [0.814, 0.961] | 0.004 | 852 / 28495 (3.0%) | 1.527 [1.349, 1.727] | <0.001 |
| West Midlands | 472 (11.1%) | 2603 / 16555 (15.7%) | 1.261 [1.168, 1.362] | <0.001 | 1077 / 26424 (4.1%) | 1.763 [1.563, 1.987] | <0.001 |
| Yorkshire & Humber | 442 (10.4%) | 1758 / 14605 (12.0%) | 0.934 [0.86, 1.014] | 0.103 | 1246 / 21543 (5.8%) | 2.481 [2.204, 2.794] | <0.001 |
| **LTCF size** |  |  |  |  |  |  |  |
| < 25 beds | 790 (18.6%) | 733 / 12214 (6.0%) | 1 |  | 371 / 17421 (2.1%) | 1 |  |
| 25-50 beds | 2204 (52%) | 6774 / 63866 (10.6%) | 1 [0.919, 1.088] | 0.991 | 3896 / 93627 (4.2%) | 1.045 [0.934, 1.169] | 0.445 |
| > 50 beds | 1244 (29.4%) | 9476 / 64300 (14.7%) | 1.028 [0.943, 1.12] | 0.532 | 4997 / 101316 (4.9%) | 0.949 [0.847, 1.062] | 0.361 |
| **Quality rating - leadership** |  |  |  |  |  |  |  |
| Good or outstanding | 3100 (73.1%) | 12240 / 104345 (11.7%) | 1 |  | 7034 / 156938 (4.5%) | 1 |  |
| Inadequate / requires improvement | 1074 (25.3%) | 4567 / 34428 (13.3%) | 1.009 [0.97, 1.049] | 0.659 | 2108 / 52190 (4.0%) | 0.786 [0.746, 0.828] | <0.001 |
| No rating | 64 (1.5%) | 176 / 1607 (11.0%) | 0.975 [0.828, 1.149] | 0.763 | 122 / 3236 (3.8%) | 0.917 [0.761, 1.104] | 0.360 |
| **Primary type of care** |  |  |  |  |  |  |  |
| > 65 years | 3306 (78%) | 12414 / 105543 (11.8%) | 1 |  | 6563 / 159666 (4.1%) | 1 |  |
| Dementia | 932 (22%) | 4569 / 34837 (13.1%) | 0.941 [0.905, 0.978] | 0.002 | 2701 / 52698 (5.1%) | 1.058 [1.008, 1.11] | 0.022 |
| **Sickness pay** |  |  |  |  |  |  |  |
| None | 304 (7.2%) | 1501 / 10750 (14.0%) | 1 |  | 860 / 16601 (5.2%) | 1 |  |
| Statutory | 654 (15.4%) | 12563 / 106349 (11.8%) | 0.797 [0.75, 0.847] | <0.001 | 6736 / 155548 (4.3%) | 0.768 [0.712, 0.828] | <0.001 |
| Full or more than statutory | 3280 (77.4%) | 2919 / 23281 (12.5%) | 0.763 [0.709, 0.822] | <0.001 | 1668 / 40215 (4.1%) | 0.704 [0.643, 0.771] | <0.001 |
| **Employment of agency nurses or carers** |  |  |  |  |  |  |  |
| Not at all | 1935 (45.7%) | 4283 / 57948 (7.4%) | 1 |  | 2186 / 82190 (2.7%) | 1 |  |
| Few times a month | 669 (15.8%) | 3008 / 21906 (13.7%) | 1.456 [1.381, 1.536] | <0.001 | 1660 / 34183 (4.9%) | 1.426 [1.331, 1.528] | <0.001 |
| Few times a week | 804 (19%) | 4064 / 28724 (14.1%) | 1.47 [1.4, 1.544] | <0.001 | 2118 / 44773 (4.7%) | 1.367 [1.281, 1.458] | <0.001 |
| Most days everyday | 830 (19.6%) | 5628 / 31802 (17.7%) | 1.676 [1.598, 1.758] | <0.001 | 3300 / 51218 (6.4%) | 1.822 [1.714, 1.937] | <0.001 |
| **Employment of other agency staff** | 3342 (78.9%) | 11780 / 108254 (10.9%) | 1 |  | 6296 / 161231 (3.9%) | 1 |  |
| Few times a month | 304 (7.2%) | 1776 / 10380 (17.1%) | 1.278 [1.206, 1.356] | <0.001 | 966 / 16362 (5.9%) | 1.261 [1.172, 1.358] | <0.001 |
| Few times a week | 330 (7.8%) | 1809 / 11907 (15.2%) | 1.117 [1.054, 1.183] | <0.001 | 1114 / 18871 (5.9%) | 1.285 [1.199, 1.377] | <0.001 |
| Most days or everyday | 262 (6.2%) | 1618 / 9839 (16.4%) | 1.045 [0.982, 1.112] | 0.161 | 888 / 15900 (5.6%) | 1.026 [0.95, 1.109] | 0.514 |
| **Staff work across multiple locations** |  |  |  |  |  |  |  |
| Not at all | 3772 (89%) | 14837 / 124651 (11.9%) | 1 |  | 7954 / 186832 (4.3%) | 1 |  |
| Few times a month | 257 (6.1%) | 1343 / 9095 (14.8%) | 1.066 [1, 1.136] | 0.050 | 718 / 14718 (4.9%) | 1.036 [0.956, 1.124] | 0.387 |
| Few times a week | 189 (4.5%) | 721 / 6031 (12.0%) | 0.928 [0.854, 1.008] | 0.076 | 499 / 9776 (5.1%) | 1.189 [1.08, 1.308] | <0.001 |
| Most days or everyday | 20 (0.5%) | 82 / 603 (13.6%) | 1.264 [0.992, 1.612] | 0.058 | 93 / 1038 (9.0%) | 2.158 [1.728, 2.695] | <0.001 |
| **Staff care for infected & uninfected residents** |  |  |  |  |  |  |  |
| Never | 1131 (26.7%) | 3961 / 37134 (10.7%) | 1 |  | 2227 / 56439 (3.9%) | 1 |  |
| Rarely or sometimes | 1131 (26.7%) | 6487 / 41694 (15.6%) | 1.172 [1.121, 1.225] | <0.001 | 3567 / 65586 (5.4%) | 1.084 [1.025, 1.147] | 0.005 |
| Often or all the time | 899 (21.2%) | 5924 / 32280 (18.4%) | 1.367 [1.306, 1.432] | <0.001 | 3227 / 50003 (6.5%) | 1.164 [1.098, 1.234] | <0.001 |
| Not applicable | 1077 (25.4%) | 611 / 29272 (2.1%) | 0.338 [0.309, 0.37] | <0.001 | 243 / 40336 (0.6%) | 0.256 [0.223, 0.293] | <0.001 |
| **Cleaning frequency – communal areas** |  |  |  |  |  |  |  |
| At least twice a day | 3179 (75%) | 12559 / 103980 (12.1%) | 1 |  | 6687 / 156233 (4.3%) | 1 |  |
| Once a day | 958 (22.6%) | 4035 / 32587 (12.4%) | 1.042 [0.998, 1.089] | 0.063 | 2337 / 50197 (4.7%) | 1.104 [1.046, 1.166] | <0.001 |
| Other | 101 (2.4%) | 389 / 3813 (10.2%) | 0.702 [0.615, 0.802] | <0.001 | 240 / 5934 (4.0%) | 1.068 [0.907, 1.258] | 0.430 |
| **Cleaning frequency – communal touchpoints** |  |  |  |  |  |  |  |
| At least twice a day | 3705 (87.4%) | 15165 / 122774 (12.4%) | 1 |  | 8241 / 186384 (4.4%) | 1 |  |
| Once a day | 380 (9%) | 1117 / 11820 (9.5%) | 0.827 [0.77, 0.887] | <0.001 | 648 / 17150 (3.8%) | 0.932 [0.853, 1.018] | 0.119 |
| Other | 153 (3.6%) | 701 / 5786 (12.1%) | 1.022 [0.923, 1.132] | 0.673 | 375 / 8830 (4.2%) | 0.921 [0.806, 1.052] | 0.226 |
| **Cleaning frequency – staff rooms** |  |  |  |  |  |  |  |
| At least twice a day | 2182 (51.5%) | 8551 / 71292 (12.0%) | 1 |  | 4757 / 108952 (4.4%) | 1 |  |
| Once a day | 1676 (39.5%) | 7055 / 58451 (12.1%) | 0.928 [0.894, 0.965] | <0.001 | 3874 / 87498 (4.4%) | 0.931 [0.887, 0.977] | 0.004 |
| Other | 380 (9%) | 1377 / 10637 (12.9%) | 1.166 [1.089, 1.249] | <0.001 | 633 / 15914 (4.0%) | 0.856 [0.781, 0.939] | 0.001 |
| **PPE use** |  |  |  |  |  |  |  |
| All the time | 2952 (69.7%) | 12670 / 98867 (12.8%) | 1 |  | 6928 / 150156 (4.6%) | 1 |  |
| Direct care - all residents | 533 (12.6%) | 1557 / 16196 (9.6%) | 0.896 [0.845, 0.95] | <0.001 | 796 / 23939 (3.3%) | 0.926 [0.857, 1] | 0.049 |
| Direct care - infected residents | 62 (1.5%) | 193 / 2123 (9.1%) | 0.745 [0.639, 0.869] | <0.001 | 82 / 3073 (2.7%) | 0.616 [0.493, 0.771] | <0.001 |
| Any contact - all residents | 627 (14.8%) | 2257 / 20837 (10.8%) | 0.826 [0.786, 0.868] | <0.001 | 1336 / 31733 (4.2%) | 0.888 [0.835, 0.944] | <0.001 |
| Any contact - infected residents | 64 (1.5%) | 306 / 2357 (13.0%) | 1.111 [0.979, 1.261] | 0.104 | 122 / 3463 (3.5%) | 0.866 [0.719, 1.043] | 0.129 |
| **Barrier nursing – infected residents** |  |  |  |  |  |  |  |
| Not used | 1062 (25.1%) | 1230 / 44503 (2.8%) | 1 |  | 677 / 61683 (1.1%) | 1 |  |
| Used | 2636 (62.2%) | 15753 / 95877 (16.4%) | 3.404 [3.192, 3.63] | <0.001 | 8587 / 150681 (5.7%) | 2.776 [2.549, 3.024] | <0.001 |
| **Barrier nursing – all residents** |  |  |  |  |  |  |  |
| Not used | 1942 (45.8%) | 5233 / 61876 (8.5%) | 1 |  | 2737 / 91971 (3.0%) | 1 |  |
| Used | 2296 (54.2%) | 11750 / 78504 (15.0%) | 1.474 [1.421, 1.528] | <0.001 | 6527 / 120393 (5.4%) | 1.476 [1.408, 1.547] | <0.001 |
| **Unable to isolate residents due to non-compliance** |  |  |  |  |  |  |  |
| No | 2832 (66.8%) | 7670 / 86603 (8.9%) | 1 |  | 3939 / 128990 (3.1%) | 1 |  |
| Yes | 1406 (33.2%) | 9313 / 53777 (17.3%) | 1.346 [1.3, 1.394] | <0.001 | 5325 / 83374 (6.4%) | 1.426 [1.363, 1.492] | <0.001 |
| **Admission to the LTCF** |  |  |  |  |  |  |  |
| Baseline |  |  | 1 |  |  | 1 |  |
| Each additional admission | 4238 (100%) | 16983 / 140380 (12.1%) | 1.001 [0.999, 1.003] | 0.425 | 9264 / 212364 (4.4%) | 0.993 [0.989, 0.996] | <0.001 |
| **Week of closure to visitors** |  |  |  |  |  |  |  |
| Baseline (March 1^st^) |  |  | 1 |  |  | 1 |  |
| No. of weeks since baseline | 4238 (100%) | 16983 / 140380 (12.1%) | 1.022 [1.008, 1.036] | 0.002 | 9264 / 212364 (4.4%) | 1.016 [0.999, 1.034] | 0.068 |

Note – Both models are based on 4238 care homes due to missing data in variables (R² = 0.26).

**eTable4: Multivariable analysis of risk factors for infection in residents and staff (using the linked test-survey dataset) with multiple imputation**

| **Risk Factor** | **Residents** | | | | **Staff** | | | |
| --- | --- | --- | --- | --- | --- | --- | --- | --- |
|  | **Number per category (%)** | **Proportion with infection (%)** | **Adjusted OR (95% CI)** | **p-value** | **Number per category (%)** | **Proportion with infection (%)** | **Adjusted OR (95% CI)** | **p-value** |
| **Age^[3]^** |  |  |  |  |  |  |  |  |
| Baseline age-group |  |  | 1 |  |  |  | 1 |  |
| age | 50425 (100%) | 1401 / 50425 (2.8%) | 1.013 [1.005, 1.021] | 0.002 | 43672 (100%) | 287 / 43672 (0.7%) | 1 [0.99, 1.009] | 0.917 |
| **Gender** |  |  |  |  |  |  |  |  |
| Female | 364466 (72.3%) | 1015 / 36466 (2.8%) | 1 |  | 36556 (83.7%) | 239 / 36556 (0.7%) | 1 |  |
| Male | 13959 (27.7%) | 386 / 13959 (2.8%) | 1.041 [0.907, 1.194] | 0.568 | 7116 (16.3%) | 48 / 7116 (0.7%) | 0.954 [0.689, 1.321] | 0.777 |
| **Social deprivation** |  |  |  |  |  |  |  |  |
| All other groups | 1746 (86.1%) | 1136 / 43005 (2.6%) | 1 |  | 1237 (90.3%) | 258 / 39025 (0.7%) | 1 |  |
| Most deprived quintile | 283 (13.9%) | 265 / 7420 (3.6%) | 1.062 [0.735, 1.535] | 0.749 | 133 (9.7%) | 29 / 4647 (0.6%) | 0.893 [0.422, 1.891] | 0.767 |
| **Care sector** |  |  |  |  |  |  |  |  |
| Not for profit | 339 (16.7%) | 309 / 9437 (3.3%) | 1 |  | 205 (15.0%) | 42 / 8176 (0.5%) | 1 |  |
| For profit | 1690 (83.3%) | 1092 / 40988 (2.7%) | 1.341 [0.89, 2.02] | 0.161 | 1165 (85.0%) | 245 / 35496 (0.7%) | 1.005 [0.49, 2.06] | 0.990 |
| **Number of LTCFs in chain** |  |  |  |  |  |  |  |  |
| Single | 1009 (49.7%) | 473 / 22049 (2.1%) | 1 |  | 750 (54.7%) | 136 / 21535 (0.6%) | 1 |  |
| 2-9 LTCFs | 531 (26.2%) | 310 / 13304 (2.3%) | 1.025 [0.743, 1.413] | 0.880 | 370 (27.0%) | 86 / 12463 (0.7%) | 0.786 [0.461, 1.341] | 0.377 |
| 10+ LTCFs | 489 (24.1%) | 618 / 15072 (4.1%) | 1.55 [1.1, 2.185] | 0.012 | 250 (18.2%) | 65 / 9674 (0.7%) | 0.775 [0.402, 1.493] | 0.446 |
| **Staff to bed ratio** |  |  |  |  |  |  |  |  |
| Baseline group |  |  | 1 |  |  |  | 1 |  |
| Each one unit increased in staff : bed ratio | 2029 (100%) | 1401 / 50425 (2.8%) | 0.706 [0.474, 1.052] | 0.087 | 370 (27.0%) | 287 / 43672 (0.7%) | 0.868 [0.492, 1.53] | 0.624 |
| **Region** |  |  |  |  |  |  |  |  |
| London | 114 (5.6%) | 61 / 3184 (1.9%) | 1 |  | 93 (6.8%) | 11 / 2957 (0.4%) | 1 |  |
| East Midlands | 203 (10%) | 113 / 4743 (2.4%) | 1.325 [0.676, 2.596] | 0.412 | 148 (10.8%) | 28 / 4636 (0.6%) | 1.554 [0.519, 4.656] | 0.431 |
| East of England | 252 (12.4%) | 208 / 6672 (3.1%) | 1.523 [0.805, 2.881] | 0.196 | 224 (16.4%) | 56 / 7114 (0.8%) | 1.618 [0.582, 4.501] | 0.357 |
| North East | 84 (4.1%) | 114 / 2147 (5.3%) | 2.718 [1.294, 5.712] | 0.008 | 12 (0.9%) | 2 / 316 (0.6%) | 0.935 [0.085, 10.344] | 0.957 |
| North West | 217 (10.7%) | 191 / 5785 (3.3%) | 1.778 [0.938, 3.37] | 0.078 | 30 (2.2%) | 18 / 981 (1.8%) | 3.774 [0.918, 15.518] | 0.066 |
| South East | 454 (22.4%) | 260 / 10565 (2.5%) | 1.371 [0.757, 2.484] | 0.298 | 395 (28.8%) | 91 / 12670 (0.7%) | 1.576 [0.606, 4.103] | 0.351 |
| South West | 337 (16.6%) | 107 / 8098 (1.3%) | 0.683 [0.352, 1.326] | 0.260 | 280 (20.4%) | 22 / 8952 (0.2%) | 0.559 [0.184, 1.692] | 0.303 |
| West Midlands | 204 (10.1%) | 120 / 5250 (2.3%) | 1.181 [0.606, 2.302] | 0.626 | 142 (10.4%) | 30 / 4727 (0.6%) | 1.407 [0.461, 4.299] | 0.549 |
| Yorkshire & Humber | 164 (8.1%) | 227 / 3981 (5.7%) | 2.903 [1.502, 5.612] | 0.002 | 46 (3.4%) | 29 / 1319 (2.2%) | 5.82 [1.626, 20.834] | 0.007 |
| **LTCF size** |  |  |  |  |  |  |  |  |
| < 25 | 341 (16.8%) | 53 / 4288 (1.2%) | 1 |  | 276 (20.1%) | 21 / 4642 (0.5%) | 1 |  |
| 25 - 50 | 1023 (50.4%) | 551 / 22980 (2.4%) | 1.03 [0.614, 1.726] | 0.911 | 696 (50.8%) | 104 / 20437 (0.5%) | 0.751 [0.357, 1.581] | 0.451 |
| > 50 | 665 (32.8%) | 797 / 23157 (3.4%) | 1.232 [0.72, 2.108] | 0.447 | 398 (29.1%) | 162 / 18593 (0.9%) | 1.079 [0.489, 2.381] | 0.850 |
| **Quality rating - leadership** |  |  |  |  |  |  |  |  |
| Outstanding or good | 1480 (72.9%) | 992 / 37891 (2.6%) | 1 |  | 985 (71.9%) | 198 / 32347 (0.6%) | 1 |  |
| Inadequate or requires improvement | 521 (25.7%) | 394 / 12009 (3.3%) | 1.168 [0.869, 1.57] | 0.302 | 364 (26.6%) | 85 / 10735 (0.8%) | 1.066 [0.645, 1.759] | 0.804 |
| No rating | 28 (1.4%) | 15 / 525 (2.9%) | 0.909 [0.289, 2.858] | 0.870 | 21 (1.5%) | 4 / 590 (0.7%) | 1.031 [0.181, 5.859] | 0.973 |
| **Primary type of care** |  |  |  |  |  |  |  |  |
| > 65 years | 1599 (78.8%) | 1089 / 38965 (2.8%) | 1 |  | 1071 (78.2%) | 207 / 33268 (0.6%) | 1 |  |
| Dementia | 430 (21.2%) | 312 / 11460 (2.7%) | 0.916 [0.672, 1.248] | 0.577 | 299 (21.8%) | 80 / 10404 (0.8%) | 0.934 [0.549, 1.589] | 0.801 |
| **Payment of sickness pay** |  |  |  |  |  |  |  |  |
| None | 143 (7%) | 144 / 3750 (3.8%) | 1 |  | 98 (7.2%) | 19 / 3180 (0.6%) | 1 |  |
| Statutory sick pay | 1571 (77.4%) | 1042 / 38169 (2.7%) | 0.727 [0.453, 1.167] | 0.187 | 1067 (77.9%) | 229 / 32585 (0.7%) | 1.196 [0.5, 2.858] | 0.688 |
| Full or more than statutory | 315 (15.5%) | 215 / 8506 (2.5%) | 0.682 [0.386, 1.205] | 0.187 | 205 (15.0%) | 39 / 7907 (0.5%) | 0.784 [0.274, 2.248] | 0.651 |
| **Employment of agency nurses or carers** |  |  |  |  |  |  |  |  |
| Not employed | 929 (45.8%) | 386 / 21099 (1.8%) | 1 |  | 647 (47.2%) | 96 / 18217 (0.5%) | 1 |  |
| Employed | 1100 (54.2%) | 1015 / 29326 (3.5%) | 1.278 [0.952, 1.714] | 0.102 | 723 (52.8%) | 191 / 25455 (0.8%) | 1.241 [0.749, 2.059] | 0.402 |
| **Employment of agency “other” staff** |  |  |  |  |  |  |  |  |
| Not employed | 1582 (78%) | 987 / 38738 (2.5%) | 1 |  | 1071 (78.2%) | 201 / 33405 (0.6%) | 1 |  |
| Employed | 447 (22%) | 414 / 11687 (3.5%) | 1.278 [0.946, 1.728] | 0.110 | 299 (21.8%) | 86 / 10267 (0.8%) | 1.108 [0.654, 1.878] | 0.704 |
| **Staff work across multiple locations** |  |  |  |  |  |  |  |  |
| No | 1811 (89.3%) | 1252 / 44819 (2.8%) | 1 |  | 1212 (88.5%) | 244 / 38570 (0.6%) | 1 |  |
| Yes | 218 (10.7%) | 149 / 5606 (2.7%) | 1.006 [0.673, 1.503] | 0.977 | 158 (11.5%) | 43 / 5102 (0.8%) | 1.115 [0.578, 2.149] | 0.746 |
| **Staff care for infected & uninfected residents** |  |  |  |  |  |  |  |  |
| Never | 553 (27.3%) | 346 / 14039 (2.5%) | 1 |  | 378 (27.6%) | 96 / 12067 (0.8%) | 1 |  |
| Rarely or sometimes | 561 (27.6%) | 516 / 15443 (3.3%) | 1.331 [0.956, 1.852] | 0.091 | 362 (26.4%) | 78 / 13176 (0.6%) | 0.797 [0.449, 1.415] | 0.438 |
| Often or all the time | 428 (21.1%) | 523 / 10988 (4.8%) | 2.046 [1.447, 2.893] | <0.001 | 252 (18.4%) | 92 / 8071 (1.1%) | 1.085 [0.586, 2.008] | 0.795 |
| Not applicable | 487 (24%) | 16 / 9955 (0.2%) | 0.181 [0.094, 0.346] | <0.001 | 378 (27.6%) | 21 / 10358 (0.2%) | 0.407 [0.188, 0.881] | 0.022 |
| **Cleaning frequency – communal areas** |  |  |  |  |  |  |  |  |
| At least twice a day | 1503 (74.1%) | 1009 / 37222 (2.7%) | 1 |  | 1014 (74.0%) | 202 / 31391 (0.6%) | 1 |  |
| Other | 526 (25.9%) | 392 / 13203 (3.0%) | 1.084 [0.782, 1.503] | 0.628 | 356 (26.0%) | 85 / 12281 (0.7%) | 0.822 [0.457, 1.479] | 0.514 |
| **Cleaning frequency - communal touchpoints** |  |  |  |  |  |  |  |  |
| At least twice a day | 1767 (87.1%) | 1236 / 43685 (2.8%) | 1 |  | 1202 (87.7%) | 253 / 38457 (0.7%) | 1 |  |
| Other | 262 (12.9%) | 165 / 6740 (2.4%) | 0.946 [0.622, 1.438] | 0.794 | 168 (12.3%) | 34 / 5215 (0.7%) | 0.996 [0.454, 2.184] | 0.992 |
| **Cleaning frequency – staff rooms** |  |  |  |  |  |  |  |  |
| At least twice a day | 1049 (51.7%) | 726 / 26256 (2.8%) | 1 |  | 721 (52.6%) | 149 / 22574 (0.7%) | 1 |  |
| Other | 980 (48.3%) | 675 / 24169 (2.8%) | 0.945 [0.712, 1.253] | 0.692 | 649 (47.4%) | 138 / 21098 (0.7%) | 0.81 [0.494, 1.328] | 0.403 |
| **Use of PPE** |  |  |  |  |  |  |  |  |
| All the time | 1402 (69.1%) | 999 / 35077 (2.8%) | 1 |  | 922 (67.3%) | 203 / 29386 (0.7%) | 1 |  |
| Providing direct care | 294 (14.5%) | 221 / 7251 (3.0%) | 0.869 [0.587, 1.286] | 0.482 | 199 (14.5%) | 45 / 6409 (0.7%) | 0.91 [0.466, 1.778] | 0.782 |
| Any contact with residents | 333 (16.4%) | 181 / 8097 (2.2%) | 0.608 [0.42, 0.882] | 0.009 | 249 (18.2%) | 39 / 7877 (0.5%) | 0.782 [0.424, 1.442] | 0.432 |
| **Barrier nursing - infected residents** |  |  |  |  |  |  |  |  |
| No | 751 (37%) | 115 / 16116 (0.7%) | 1 |  | 550 (40.1%) | 61 / 15236 (0.4%) | 1 |  |
| Yes | 1278 (63%) | 1286 / 34309 (3.7%) | 3.843 [2.58, 5.726] | <0.001 | 820 (59.9%) | 226 / 28436 (0.8%) | 1.744 [0.95, 3.202] | 0.073 |
| **Barrier nursing – all residents** |  |  |  |  |  |  |  |  |
| No | 915 (45.1%) | 513 / 22537 (2.3%) | 1 |  | 631 (46.1%) | 123 / 20006 (0.6%) | 1 |  |
| Yes | 1114 (54.9%) | 888 / 27888 (3.2%) | 1.341 [1.027, 1.751] | 0.031 | 739 (53.9%) | 164 / 23666 (0.7%) | 0.748 [0.474, 1.181] | 0.213 |
| **Unable to isolate residents due to non-compliance** |  |  |  |  |  |  |  |  |
| No | 1352 (66.6%) | 636 / 32004 (2.0%) | 1 |  | 959 (70.0%) | 147 / 28660 (0.5%) | 1 |  |
| Yes | 677 (33.4%) | 765 / 18421 (4.2%) | 1.402 [1.071, 1.834] | 0.014 | 411 (30.0%) | 140 / 15012 (0.9%) | 1.611 [0.998, 2.601] | 0.051 |
| **Number of new admissions to LTCF** |  |  |  |  |  |  |  |  |
| Baseline |  |  | 1 |  |  |  | 1 |  |
| Each additional admission | 2029 (100%) | 1401 / 50425 (2.8%) | 1.006 [0.986, 1.026] | 0.549 | 1370 (100%) | 287 / 43672 (0.7%) | 1.021 [0.987, 1.056] | 0.231 |
| **Week of closure to visitors** |  |  |  |  |  |  |  |  |
| Baseline (March 1^st^) |  |  | 1 |  |  |  | 1 |  |
| Each week since March 1st | 2029 (100%) | 1404 / 50428 (2.8%) | 1.014 [0.914, 1.125] | 0.789 | 1370 (100%) | 287 / 43672 (0.7%) | 1.085 [0.918, 1.282] | 0.340 |

Note – The resident model is based on 50425 residents in 2029 care homes (intercept = -7.93, SD = 1.72). Marginal R² = 0.06. Conditional R² = 0.14. The staff model is based on 43672 staff in 1370 care homes (intercept = -7.50, SD = 2.12). Marginal R² = 0.04. Conditional R² = 0.20.

**eTable5: Multivariable analysis of risk factors for large outbreaks in staff or residents after multiple imputation**

| **Risk Factor** | **LTCFs with ≥1 cases versus LTCF with zero cases** | | | | **Large versus small outbreaks** | | | |
| --- | --- | --- | --- | --- | --- | --- | --- | --- |
|  | **Number of LTCFs (%)** | **Proportion with infection (%)** | **Adjusted OR (95% CI)** | **p-value** | **Number of LTCFs (%)** | **Proportion with infection (%)** | **Adjusted OR (95% CI)** | **p-value** |
| **Social deprivation** |  |  |  |  |  |  |  |  |
| All other quintiles | 3505 (82.7%) | 1948 / 3505 (55.6%) | 1 |  | 1948 (81.2%) | 324 / 1948 (16.6%) | 1 |  |
| Most deprived quintile | 733 (17.3%) | 452 / 733 (61.7%) | 1.077 [0.848, 1.368] | 0.542 | 452 (18.8%) | 98 / 452 (21.7%) | 1.286 [0.964, 1.714] | 0.087 |
| **Care sector** |  |  |  |  |  |  |  |  |
| Not for profit | 714 (16.8%) | 455 / 714 (63.7%) | 1 |  | 455 (19%) | 67 / 455 (14.7%) | 1 |  |
| For profit | 3524 (83.2%) | 1945 / 3524 (55.2%) | 1.16 [0.88, 1.528] | 0.292 | 1945 (81%) | 355 / 1945 (18.3%) | 1.723 [1.212, 2.45] | 0.002 |
| **Number of LTCFs in chain** |  |  |  |  |  |  |  |  |
| Single provider | 2054 (48.5%) | 995 / 2054 (48.4%) | 1 |  | 995 (41.5%) | 154 / 995 (15.5%) | 1 |  |
| 2-9 LTCFs | 1227 (29%) | 718 / 1227 (58.5%) | 0.977 [0.8, 1.193] | 0.819 | 718 (29.9%) | 123 / 718 (17.1%) | 0.976 [0.741, 1.286] | 0.864 |
| 10+ LTCFs | 957 (22.6%) | 687 / 957 (71.8%) | 1.282 [1.004, 1.636] | 0.046 | 687 (28.6%) | 145 / 687 (21.1%) | 1.314 [0.987, 1.75] | 0.062 |
| **Staff to bed ratio** |  |  |  |  |  |  |  |  |
| Baseline |  |  | 1 |  |  |  | 1 |  |
| Each one unit increase in staff : bed ratio | 4238 (100%) | 2400 / 4238 (56.6%) | 1.134 [0.937, 1.373] | 0.196 | 2400 (100%) | 422 / 2400 (17.6%) | 1.082 [0.889, 1.315] | 0.432 |
| **Region** |  |  |  |  |  |  |  |  |
| London | 229 (5.4%) | 186 / 229 (81.2%) | 1 |  | 186 (7.8%) | 18 / 186 (9.7%) | 1 |  |
| East Midlands | 406 (9.6%) | 219 / 406 (53.9%) | 0.261 [0.157, 0.436] | <0.001 | 219 (9.1%) | 31 / 219 (14.2%) | 1.766 [0.932, 3.343] | 0.081 |
| East of England | 470 (11.1%) | 247 / 470 (52.6%) | 0.231 [0.14, 0.38] | <0.001 | 247 (10.3%) | 43 / 247 (17.4%) | 2.06 [1.121, 3.785] | 0.020 |
| North East | 248 (5.9%) | 168 / 248 (67.7%) | 0.314 [0.177, 0.557] | <0.001 | 168 (7%) | 45 / 168 (26.8%) | 3.279 [1.739, 6.182] | <0.001 |
| North West | 596 (14.1%) | 371 / 596 (62.2%) | 0.348 [0.211, 0.573] | <0.001 | 371 (15.5%) | 63 / 371 (17.0%) | 1.905 [1.059, 3.425] | 0.031 |
| South East | 786 (18.5%) | 430 / 786 (54.7%) | 0.271 [0.168, 0.437] | <0.001 | 430 (17.9%) | 68 / 430 (15.8%) | 1.878 [1.062, 3.323] | 0.030 |
| South West | 589 (13.9%) | 201 / 589 (34.1%) | 0.104 [0.064, 0.171] | <0.001 | 201 (8.4%) | 45 / 201 (22.4%) | 2.881 [1.564, 5.306] | 0.001 |
| West Midlands | 472 (11.1%) | 315 / 472 (66.7%) | 0.352 [0.211, 0.586] | <0.001 | 315 (13.1%) | 63 / 315 (20.0%) | 2.587 [1.443, 4.636] | 0.001 |
| Yorkshire & Humber | 442 (10.4%) | 263 / 442 (59.5%) | 0.29 [0.174, 0.482] | <0.001 | 263 (11%) | 46 / 263 (17.5%) | 2.016 [1.099, 3.698] | 0.024 |
| **LTCF size** |  |  |  |  |  |  |  |  |
| < 25 beds | 790 (18.6%) | 215 / 790 (27.2%) | 1 |  | 215 (9%) | 28 / 215 (13.0%) | 1 |  |
| 25-50 beds | 2204 (52%) | 1206 / 2204 (54.7%) | 1.699 [1.335, 2.161] | <0.001 | 1206 (50.3%) | 165 / 1206 (13.7%) | 0.795 [0.5, 1.264] | 0.333 |
| > 50 beds | 1244 (29.4%) | 979 / 1244 (78.7%) | 2.981 [2.231, 3.984] | <0.001 | 979 (40.8%) | 229 / 979 (23.4%) | 1.463 [0.911, 2.349] | 0.115 |
| **Quality rating - leadership** |  |  |  |  |  |  |  |  |
| Well led or good | 3100 (73.1%) | 1709 / 3100 (55.1%) | 1 |  | 1709 (71.2%) | 312 / 1709 (18.3%) | 1 |  |
| Inadequate or requires improvement | 1074 (25.3%) | 653 / 1074 (60.8%) | 1.155 [0.945, 1.411] | 0.159 | 653 (27.2%) | 107 / 653 (16.4%) | 0.822 [0.636, 1.064] | 0.137 |
| No rating | 64 (1.5%) | 38 / 64 (59.4%) | 0.987 [0.494, 1.971] | 0.969 | 38 (1.6%) | 3 / 38 (7.9%) | 0.376 [0.112, 1.261] | 0.113 |
| **Primary type of LTCF** |  |  |  |  |  |  |  |  |
| > 65 years | 3306 (78%) | 1808 / 3306 (54.7%) | 1 |  | 1808 (75.3%) | 305 / 1808 (16.9%) | 1 |  |
| Dementia | 932 (22%) | 592 / 932 (63.5%) | 1.045 [0.847, 1.29] | 0.680 | 592 (24.7%) | 117 / 592 (19.8%) | 0.981 [0.762, 1.263] | 0.880 |
| **Sickness Pay** |  |  |  |  |  |  |  |  |
| None | 304 (7.2%) | 182 / 304 (59.9%) | 1 |  | 182 (7.6%) | 41 / 182 (22.5%) | 1 |  |
| Statutory sickness pay | 654 (15.4%) | 1796 / 3280 (54.8%) | 0.822 [0.592, 1.142] | 0.242 | 1796 (74.8%) | 310 / 1796 (17.3%) | 0.671 [0.455, 0.99] | 0.044 |
| Full or more than statutory | 3280 (77.4%) | 422 / 654 (64.5%) | 0.861 [0.576, 1.287] | 0.465 | 422 (17.6%) | 71 / 422 (16.8%) | 0.702 [0.434, 1.134] | 0.148 |
| **Employment of agency nurses or carers** |  |  |  |  |  |  |  |  |
| Not at all | 1935 (45.7%) | 784 / 1935 (40.5%) | 1 |  | 784 (32.7%) | 92 / 784 (11.7%) | 1 |  |
| Few times a month | 669 (15.8%) | 534 / 804 (66.4%) | 1.511 [1.178, 1.939] | 0.001 | 419 (17.5%) | 79 / 419 (18.9%) | 1.634 [1.15, 2.322] | 0.006 |
| Few times a week | 804 (19%) | 419 / 669 (62.6%) | 1.571 [1.239, 1.991] | <0.001 | 534 (22.3%) | 97 / 534 (18.2%) | 1.503 [1.077, 2.097] | 0.016 |
| Most days or everyday | 830 (19.6%) | 663 / 830 (79.9%) | 2.414 [1.855, 3.14] | <0.001 | 663 (27.6%) | 154 / 663 (23.2%) | 2.191 [1.598, 3.003] | <0.001 |
| **Employment of agency “other” staff** |  |  |  |  |  |  |  |  |
| Not at all | 3342 (78.9%) | 1736 / 3342 (51.9%) | 1 |  | 1736 (72.3%) | 292 / 1736 (16.8%) | 1 |  |
| Few times a month | 304 (7.2%) | 222 / 304 (73.0%) | 1.477 [1.031, 2.116] | 0.033 | 222 (9.3%) | 46 / 222 (20.7%) | 1.188 [0.817, 1.728] | 0.366 |
| Few times a week | 330 (7.8%) | 225 / 330 (68.2%) | 1.27 [0.912, 1.77] | 0.158 | 225 (9.4%) | 50 / 225 (22.2%) | 1.241 [0.86, 1.792] | 0.248 |
| Most days or everyday | 262 (6.2%) | 217 / 262 (82.8%) | 1.89 [1.225, 2.917] | 0.004 | 217 (9%) | 34 / 217 (15.7%) | 0.679 [0.445, 1.034] | 0.071 |
| **Staff work across multiple sites** |  |  |  |  |  |  |  |  |
| Not at all | 3772 (89%) | 2093 / 3772 (55.5%) | 1 |  | 2093 (87.2%) | 367 / 2093 (17.5%) | 1 |  |
| Few times a month | 257 (6.1%) | 170 / 257 (66.1%) | 0.927 [0.64, 1.34] | 0.685 | 170 (7.1%) | 33 / 170 (19.4%) | 1.082 [0.71, 1.648] | 0.714 |
| Few times a week | 189 (4.5%) | 122 / 189 (64.6%) | 1.159 [0.763, 1.761] | 0.490 | 122 (5.1%) | 19 / 122 (15.6%) | 0.943 [0.555, 1.601] | 0.827 |
| Most days or everyday | 20 (0.5%) | 15 / 20 (75.0%) | 1.414 [0.458, 4.372] | 0.547 | 15 (0.6%) | 3 / 15 (20.0%) | 1.492 [0.383, 5.81] | 0.564 |
| **Staff care for infected & uninfected residents** |  |  |  |  |  |  |  |  |
| Never | 1131 (26.7%) | 602 / 1131 (53.2%) | 1 |  | 602 (25.1%) | 101 / 602 (16.8%) | 1 |  |
| Rarely or sometimes | 1131 (26.7%) | 871 / 1131 (77.0%) | 1.744 [1.399, 2.175] | <0.001 | 871 (36.3%) | 164 / 871 (18.8%) | 0.99 [0.74, 1.324] | 0.946 |
| Often or all of the time | 899 (21.2%) | 739 / 899 (82.2%) | 2.608 [2.03, 3.351] | <0.001 | 739 (30.8%) | 141 / 739 (19.1%) | 1.008 [0.745, 1.364] | 0.958 |
| Not applicable | 1077 (25.4%) | 188 / 1077 (17.5%) | 0.377 [0.298, 0.478] | <0.001 | 188 (7.8%) | 16 / 188 (8.5%) | 0.6 [0.332, 1.084] | 0.090 |
| **Cleaning frequency – communal areas** |  |  |  |  |  |  |  |  |
| At least twice per day | 3179 (75%) | 1773 / 3179 (55.8%) | 1 |  | 1773 (73.9%) | 312 / 1773 (17.6%) | 1 |  |
| Once per day | 958 (22.6%) | 557 / 958 (58.1%) | 1.022 [0.811, 1.288] | 0.853 | 557 (23.2%) | 103 / 557 (18.5%) | 1.123 [0.846, 1.491] | 0.421 |
| Other | 101 (2.4%) | 70 / 101 (69.3%) | 1.28 [0.643, 2.546] | 0.482 | 70 (2.9%) | 7 / 70 (10.0%) | 0.545 [0.21, 1.415] | 0.212 |
| **Cleaning frequency – communal touchpoints** |  |  |  |  |  |  |  |  |
| At least twice per day | 3705 (87.4%) | 2109 / 3705 (56.9%) | 1 |  | 2109 (87.9%) | 385 / 2109 (18.3%) | 1 |  |
| Once per day | 380 (9%) | 187 / 380 (49.2%) | 0.798 [0.578, 1.101] | 0.170 | 187 (7.8%) | 23 / 187 (12.3%) | 0.658 [0.402, 1.078] | 0.097 |
| Other | 153 (3.6%) | 104 / 153 (68.0%) | 1.35 [0.77, 2.365] | 0.295 | 104 (4.3%) | 14 / 104 (13.5%) | 0.841 [0.414, 1.707] | 0.631 |
| **Cleaning frequency – staff rooms** |  |  |  |  |  |  |  |  |
| Twice per day | 2182 (51.5%) | 1200 / 2182 (55.0%) | 1 |  | 1200 (50%) | 215 / 1200 (17.9%) | 1 |  |
| At least once per day | 1676 (39.5%) | 1003 / 1676 (59.8%) | 0.995 [0.818, 1.212] | 0.963 | 1003 (41.8%) | 174 / 1003 (17.3%) | 0.901 [0.701, 1.157] | 0.413 |
| Other | 380 (9%) | 197 / 380 (51.8%) | 1.171 [0.844, 1.625] | 0.344 | 197 (8.2%) | 33 / 197 (16.8%) | 0.994 [0.638, 1.548] | 0.979 |
| **Use of PPE** |  |  |  |  |  |  |  |  |
| All the time | 2952 (69.7%) | 1743 / 2952 (59.0%) | 1 |  | 1743 (72.6%) | 309 / 1743 (17.7%) | 1 |  |
| Direct care – all residents | 533 (12.6%) | 250 / 533 (46.9%) | 0.917 [0.706, 1.189] | 0.512 | 250 (10.4%) | 44 / 250 (17.6%) | 1.151 [0.797, 1.661] | 0.454 |
| Direct care – infected residents | 62 (1.5%) | 33 / 62 (53.2%) | 0.704 [0.373, 1.329] | 0.279 | 33 (1.4%) | 3 / 33 (9.1%) | 0.464 [0.136, 1.584] | 0.220 |
| Any contact – all residents | 627 (14.8%) | 335 / 627 (53.4%) | 0.785 [0.618, 0.999] | 0.049 | 335 (14%) | 59 / 335 (17.6%) | 0.892 [0.645, 1.234] | 0.491 |
| Any contact – infected residents | 64 (1.5%) | 39 / 64 (60.9%) | 0.962 [0.502, 1.843] | 0.906 | 39 (1.6%) | 7 / 39 (17.9%) | 1.306 [0.547, 3.119] | 0.547 |
| **Use of barrier nursing – infected residents** |  |  |  |  |  |  |  |  |
| No | 1062 (25.1%) | 345 / 1602 (21.5%) | 1 |  | 345 (14.4%) | 37 / 345 (10.7%) | 1 |  |
| Yes | 2636 (62.2%) | 2055 / 2636 (78.0%) | 5.001 [4.166, 6.005] | <0.001 | 2055 (85.6%) | 385 / 2055 (18.7%) | 1.236 [0.827, 1.846] | 0.301 |
| **Use of barrier nursing – all residents** |  |  |  |  |  |  |  |  |
| No | 1942 (45.8%) | 889 / 1942 (45.8%) | 1 |  | 889 (37%) | 120 / 889 (13.5%) | 1 |  |
| Yes | 2296 (54.2%) | 1511 / 2296 (65.8%) | 1.657 [1.396, 1.966] | <0.001 | 1511 (63%) | 302 / 1511 (20.0%) | 1.539 [1.204, 1.966] | 0.001 |
| **Unable to isolate due to non-compliance** |  |  |  |  |  |  |  |  |
| No | 2832 (66.8%) | 1309 / 2832 (46.2%) | 1 |  | 1309 (54.5%) | 177 / 1309 (13.5%) | 1 |  |
| Yes | 1406 (33.2%) | 1091 / 1406 (77.6%) | 1.811 [1.498, 2.19] | <0.001 | 1091 (45.5%) | 245 / 1091 (22.5%) | 1.486 [1.183, 1.867] | 0.001 |
| **New admission to the LTCF** |  |  |  |  |  |  |  |  |
| Baseline |  |  | 1 |  |  |  | 1 |  |
| Each additional admission | 4238 (100%) | 2400 / 4238 (56.6%) | 1.068 [1.045, 1.092 | <0.001 | 2400 (100%) | 422 / 2400 (17.6%) | 0.989 [0.972, 1.007] | 0.227 |
| **Closure to visitors** |  |  |  |  |  |  |  |  |
| Baseline (1^st^ March 2020) |  |  | 1 |  |  |  | 1 |  |
| Each additional week since baseline | 4238 (100%) | 2400 / 4238 (56.6%) | 1.024 [0.958, 1.094] | 0.484 | 2400 (100%) | 422 / 2400 (17.6%) | 1.06 [0.973, 1.156] | 0.183 |

Note – Cases vs. no cases model is based on 4238 LTCFs, of which 2400 LTCFs had cases (R² = 0.40). Large vs. small outbreaks model is based on 2400 care homes, of which 422 LTCFs were considered to have a “large outbreak” (R² = 0.08).

1. Individual-level data presented
2. Baseline group for residents=65 years; baseline group for staff=16 years
3. 65 years for residents, 16 years for staff
